# Pre-existing antibodies targeting a linear epitope on SARS-CoV-2 S2 cross-reacted with commensal gut bacteria and shaped vaccine induced immunity

**DOI:** 10.1101/2021.07.13.21260404

**Authors:** Liqiu Jia, Shufeng Weng, Jing Wu, Xiangxiang Tian, Yifan Zhang, Xuyang Wang, Jing Wang, Dongmei Yan, Wanhai Wang, Fang Fang, Zhaoqin Zhu, Chao Qiu, Wenhong Zhang, Ying Xu, Yanmin Wan

## Abstract

The origins of pre-existing SARS-CoV-2 cross-reactive antibodies and their potential impacts on vaccine efficacy have not been fully clarified. In this study, we demonstrated that S2 was the prevailing target of the pre-existing S protein cross-reactive antibodies in both healthy human and SPF mice. A dominant antibody epitope was identified on the connector domain of S2 (1147-SFKEELDKYFKNHT-1160, P144), which could be recognized by pre-existing antibodies in both human and mouse. Through metagenomic sequencing and fecal bacteria transplant, we proved that the generation of S2 cross-reactive antibodies was associated with commensal gut bacteria. Furthermore, six P144 specific monoclonal antibodies were isolated from naïve SPF mice and proved to cross-react with commensal gut bacteria collected from both human and mouse. Mice with high levels of pre-existing S2 cross-reactive antibodies mounted higher S protein specific binding antibodies, especially against S2, after being immunized with a SARS-CoV-2 S DNA vaccine. Similarly, we found that levels of pre-existing S2 and P144 reactive antibodies correlated positively with RBD specific binding antibody titers after two doses of inactivated SARS-CoV-2 vaccination in human. Finally, we provided data demonstrating that immunization of a SARS-CoV-2 S DNA vaccine could alter the gut microbiota compositions of mice.

## Introduction

Antibodies are vital components of the immune system that mediate protection against infections (1). When confronting infections, the actual role of pre-existing antibody depends on the following features (2): High titers of broadly neutralizing antibodies can protect the host against infection. While, when the pre-existing antibodies are non-neutralizing or with only a narrow neutralizing spectrum, hosts may not be sterilely protected or only be protected against specific serotypes of viruses. In addition to defending hosts against infections, pre-existing antibodies can also impact host immune responses upon infection or vaccination (3–5), which is best exemplified by the observations showing that pre-existing antibodies shaped the recall immune responses against influenza (6, 7).

For most occasions, pre-existing antibodies in adults derive from previous infection or vaccination except some “naturally” produced, poly-reactive antibodies (2, 8). When encountering a newly emerged or mutated virus, cross-reactive antibodies induced by previously occurred, phylogenetically closely related viruses constitute the main body of the pre-existing cross-reactive antibodies. The effect of this kind of pre-existing antibodies has been extensively investigated especially for infections of influenza (3, 7, 9) and flaviviruses (10–12). Of note, previous infection by phylogenetically similar viruses is not the sole source of pre-existing cross-reactive antibodies, as it has been clearly clarified that pre-existing antibodies against HIV-1 gp41 may stem from exposures to certain commensal gut bacteria (13–15). Besides, autoimmune diseases caused by cross-reactivities between microbial and self-antigens also implied that commensal gut bacteria represent important sources of cross-reactive antibodies (16–19).

Pre-existing antibodies against SARS-CoV-2 have also been observed in uninfected healthy individuals, which are speculated to be engendered by previous exposures to human common cold coronaviruses (20–26) or SARS-CoV (27–29). Meanwhile, sequence analyses (30) and a clinical observation (31) suggest that pre-existing SARS-CoV-2 antibodies might be engendered by common human pathogens and childhood vaccination. Although these two explanations are not mutually exclusive, they both need more experimental evidence to support.

In this study, we found that higher levels of SARS-CoV-2 S2 protein specific antibodies existed in both healthy human and naïve SPF mice. To track the potential origins of these pre-existing cross-reactive antibodies, we mapped and identified a dominant linear antibody epitope on S2, which could be recognized by pre-existing antibodies from both healthy human and naïve SPF mice. Monoclonal antibodies against this linear epitope were isolated from naïve SPF mice and proved to cross-react with commensal gut bacteria collected from both healthy human and naïve SPF mouse. Moreover, despite having been discussed iteratively (32, 33), the influences of pre-existing cross-reactive immunities on COVID-19 responses have not been clarified. Here we showed that high levels of pre-existing antibodies did not impair the immunogenicity of a candidate DNA vaccine encoding SARS-CoV-2 spike protein. On the contrary, mice with high levels of pre-existing antibodies mounted stronger S2 specific binding antibody responses compared with mice with low levels of pre-existing antibodies after immunization with a candidate DNA vaccine. Meanwhile, we showed that inoculation of the SARS-CoV-2 S DNA vaccine could significantly alter the gut microbiota compositions of mice.

## Results

### Pre-existing antibodies recognizing a dominant linear epitope on SARS-CoV-2 S2 protein were detected in both human and mice

Pre-existing antibodies cross-react with SARS-CoV-2 S protein have been found in uninfected individuals by multiple previous studies (22, 25, 26, 34). It was postulated that the pre-existing immunities against SARS-CoV-2 might be induced by previous exposure to seasonal human coronaviruses (22, 32, 33, 35, 36). However, contradictive evidence suggested that human common cold coronavirus infection did not necessarily induce antibodies cross-reactive with SARS-CoV-2 spike protein (28, 37, 38). In addition to this hypothesis, an alternative explanation suggested that the cross-reactive immunities to SARS-CoV-2 might derive from other common human pathogens and vaccines (30).

To track the origins of the pre-existing cross-reactive antibodies to SARS-CoV-2 spike protein, in this study, we first measured the levels of pre-existing S protein specific antibodies in healthy human individuals and SPF mice. Our data showed that the cross-reactive antibody responses against S2 were significantly stronger than those against S1 in plasma samples of healthy human collected both pre (2016 cohort) and post (2020 cohort) the outbreak of COVID-19 pandemic (Figure 1A and 1B). More strikingly, our data showed that binding antibodies targeting S2 could also be detected in two strains of naïve SPF mice (Figure 1C and 1D). And this finding was further confirmed by Western-blotting (WB) assays, which showed that mouse sera with high OD values (Detected by ELISA) (Figure 2A) bound specifically with purified S2 while not S1 (Figure 2B). Quite interestingly, the WB results indicated that cross-reactive antibodies against S2 also existed in the serum of a mouse (#487) with no detectable ELISA binding signal (Figure 2A and 2B). We next performed linear antibody epitope mapping using an in-house developed method of peptide competition ELISA. Our data showed that a single peptide (P144, aa1145-aa1162, 18-mer) accounted for most of the observed pre-existing antibody responses towards S2 in mice (Supplementary Figure 1). Via employing a series of truncated peptides based on P144, we determined the minimal range of this epitope (1147-SFKEELDKYFKNHT-1160), which locates on the connector domain (adjacent to the N-terminal of HR2 domain) (Figure 2C). We also proved that antibodies recognizing this epitope widely existed in both healthy human and naïve SPF mice through competitive ELISA assays (Figure 3).

**Figure 1.**
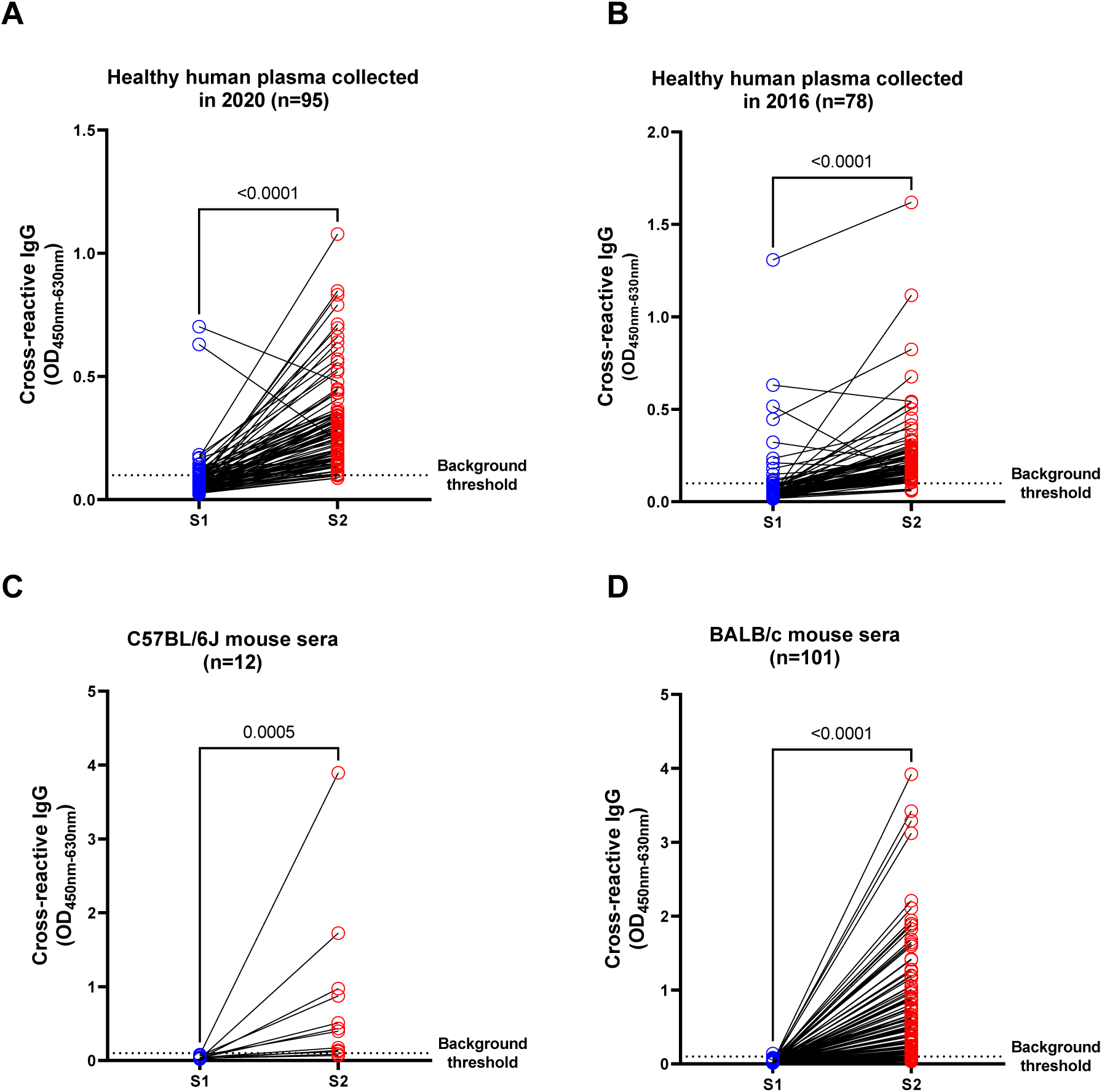
Pre-existing cross-reactive antibodies against SARS-CoV-2 S protein observed in both healthy human and naïve SPF mice predominantly targeted S2. The pre-existing cross-reactive antibodies against S1 and S2 were measured using an in-house ELISA method (Sample dilution factor: 100). (**A**) Plasma samples of healthy individuals collected in 2020 (n=95). (**B**) Plasma samples of healthy individuals collected in 2016 (n=78). (**C**) Sera of naïve C57BL/6J mice (n=12). (**D**) Sera of naïve BALB/c mice (n=101). The dotted lines show the threshold of background (3 folds of the average background OD). Statistical analyses were performed using the method of paired t-test.

**Figure 2.**
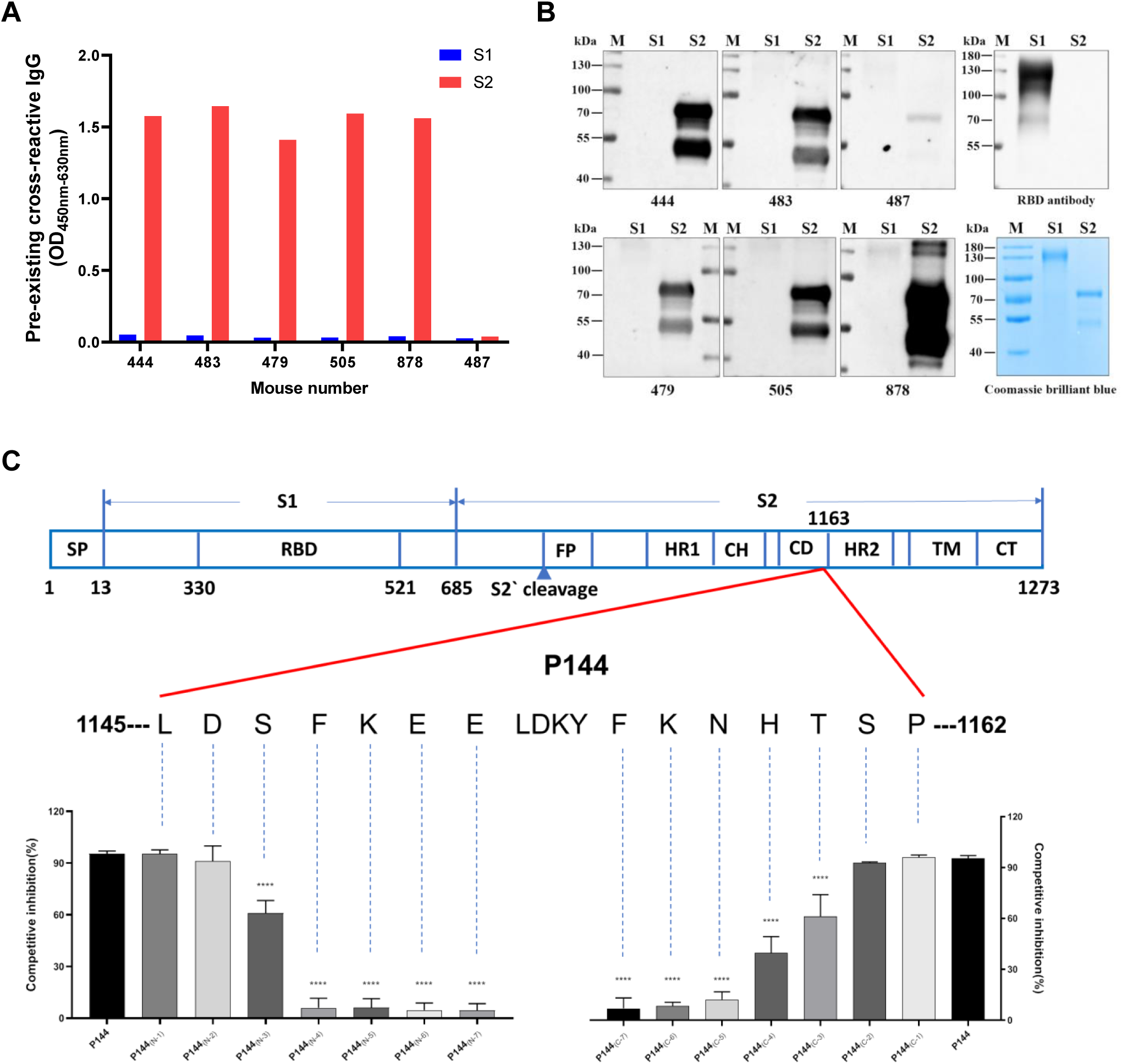
The pre-existing S protein binding antibodies in naïve SPF mice recognized S2 exclusively and a dominant linear antibody epitope was identified on the connector domain. (**A**) The pre-existing S1 and S2 reactive antibody levels in the sera of 6 representative mice. (**B**) WB assays of pre-existing cross-reactive antibodies for 6 representative mouse serum samples. The purities of S1 and S2 proteins were shown by coomassie blue staining. (**C**) The minimal epitope of P144 was defined using a method of competitive ELISA (Data shown as mean±SD, n=5). Purified S2 protein was used as the coating antigen and truncated peptides derived from P144 were used as competitors. The decreases of competitive inhibition reflected the necessity of each amino acid for the epitope recognition. Statistical differences among groups was analyzed using One-way ANOVA. ****, P<0.0001. M: molecular weight marker; SP, signal peptide; RBD, receptor-binding domain; FP, fusion peptide; HR, heptad repeat; CH, central helix; CD, connector domain; TM, transmembrane domain; CT, cytoplasmic tail.

**Figure 3.**
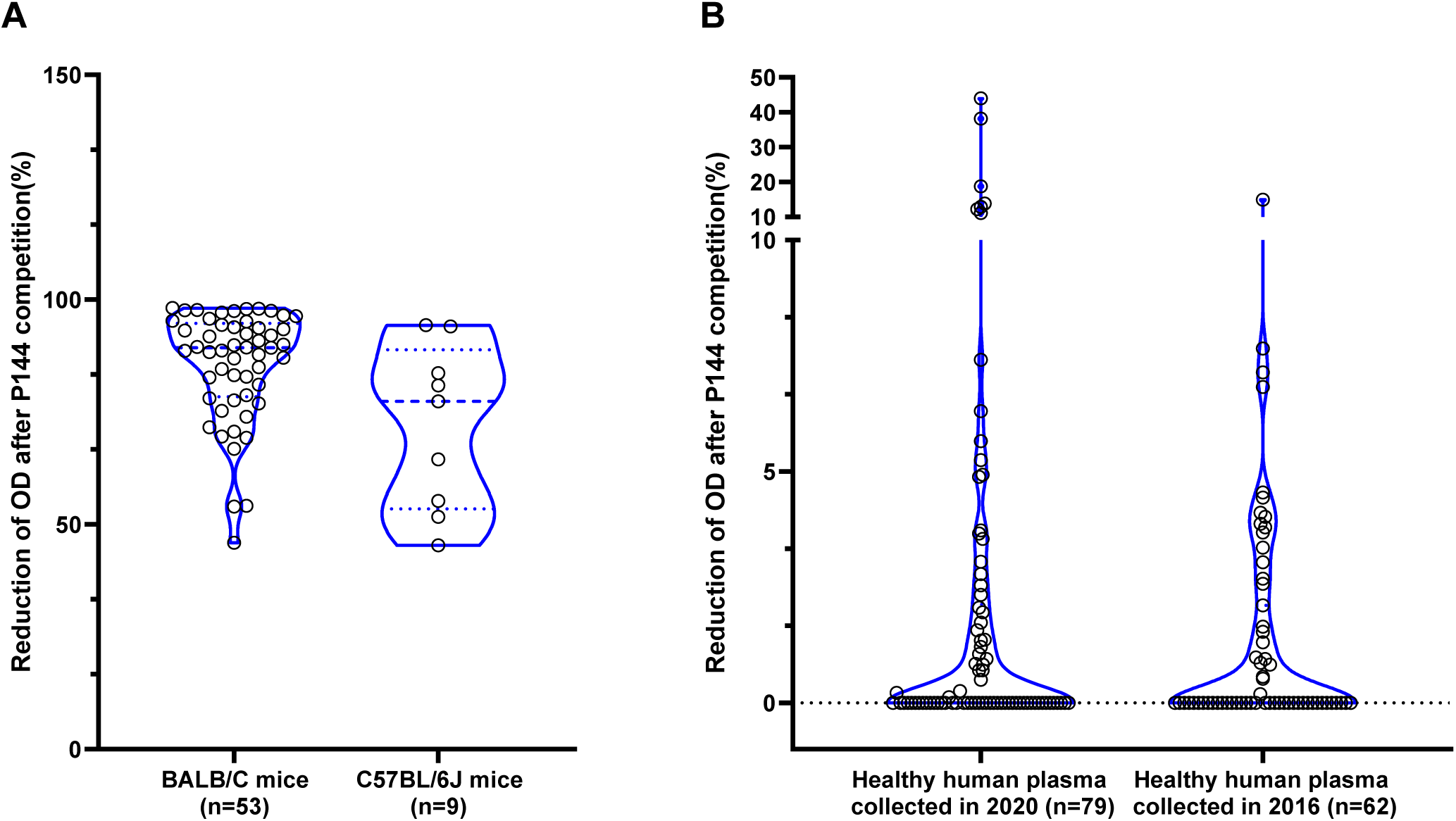
P144 reactive antibodies widely existed in both healthy human and naïve SPF mice. P144 specific binding antibodies were detected using a method of competitive ELISA. (**A**) For the detections of P144 specific binding antibodies in naïve SPF mice, purified S2 protein was used as the coating antigen and P144 peptide was used as the competitor. (**B**) For the detections of P144 specific binding antibodies in healthy individuals, purified BSA-P144 conjugate was used as the coating antigen and P144 peptide was used as the competitor. In both experiments, the reduction of OD value reflected the presence of P144 binding antibodies.

### The P144 specific antibody responses could be engendered by exposures to certain commensal gut bacteria

To explore the potential origins of the pre-existed P144 specific antibodies, we first performed phylogenetic analyses among SARS-CoV-2 and other human coronaviruses. The results showed that the aa sequence of P144 was highly conserved among SARS-CoV-2 variants and SARS-CoV, while the similarities between P144 and MERS-CoV or seasonal human coronaviruses were relatively low, especially within the range of predicted antibody binding epitope (boxed fragment) (Supplementary Figure 2). The possibility of coronavirus infection in our SPF-mouse colonies was excluded by serum screenings using commercialized mouse hepatitis virus (MHV) antigen (Cat# SBJ-M0051-48T, SinBeiJia Biological Technology Co., Ltd, China) and antibody detection kits (Cat#SY-M02196, Shanghai Shuangying Biotechnology Co., Ltd, China) (Data not shown).

Subsequently, to investigate whether environmental factors contribute to the induction of these S2 cross-reactive antibodies, we compared the levels of pre-existing S2 binding antibodies between mice housed in SPF condition and mice maintained in a sterile isolation pack. Our data showed that the levels of pre-existing S2 binding antibodies were significantly higher in SPF mice (Figure 4A). Through metagenomic sequencing, we further demonstrated that the compositions of commensal gut bacteria were significantly different between mice housed in different environments (Supplementary Figure 3A). The abundance of bacteroidaceae, prevotellaceace and parabacteroides increased significantly in the commensal gut bacteria of SPF mice (Figure 4B). Moreover, frequencies of memory B cells measured by B cell ELISPOT (Supplementary Figure 3B) and frequencies of S2 specific B cells (CD45+CD19+S2+) measured by flowcytometry (Supplementary Figure 3C) were significantly higher in mesenteric lymph nodes (MLNs) than those in spleens of mice with pre-existing S2 binding antibodies. Consistently, via 16s rDNA sequencing, we found that the gut microbiota compositions of SPF mice with different levels of pre-existing S2 binding antibodies might be different (Supplementary Figure 4). To further clarify the role of gut microbiota in the induction of S2 cross-reactive antibodies, mice fed in a sterile isolation pack were transplanted with fecal bacteria prepared from SPF mice (Figure 4C). We found that the abundances of P144 reactive antibodies in mouse sera significantly increased after fecal microbiota transplantation (FMT) (Figure 4D). These results collectively suggested that the S2 cross-reactive antibodies could be induced by exposures to certain microbial antigens.

**Figure 4.**
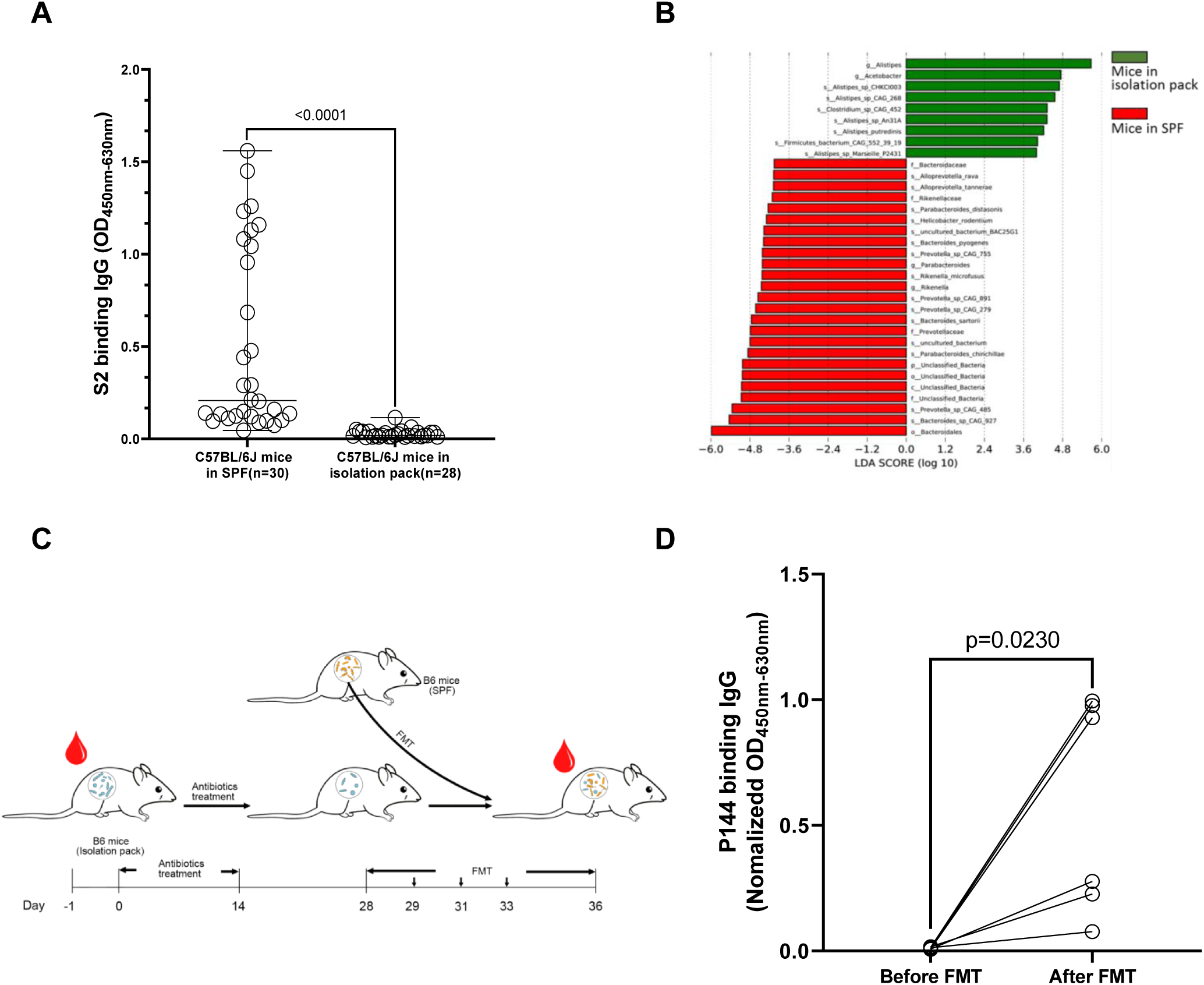
The presence of the pre-existing S2 reactive antibodies was associated with commensal gut bacteria. **(A)** Comparison of the levels of S2 reactive antibodies between naïve mice maintained under SPF condition and mice housed in a sterile isolation pack. **(B)** Metagenomic analyses of gut microbiota compositions between mice from different housing environments. Differences of bacterial abundance were manifested by linear discriminant analysis (LDA) . **(C)** Schematic overview of the fecal bacteria transplantation (n=6). The mice bred in a sterile isolation pack were treated with a mixture of ampicillin (1g/L), metronidazole (0.5g/L), vancomycin (0.5g/L) and gentamycin (0.5g/L) dissolved in drinking water supplemented with D-glucose (36.8g/L) for 14 days. Two weeks later, fecal bacteria were freshly prepared from SPF mice and delivered to antibiotic treated mice via oral gavage. **(D)** Comparison of P144 reactive antibodies in mouse sera collected before and after FMT. The OD value of P144 binding antibody was normalized to the concentration of total IgG for each mouse. Data are shown as mean±SD (n=6). Statistical analyses were performed by the method of paired t-test. (p), Phylum. (c), Class. (o), Order. (f), Family. (g), Genus. (s), Species.

### P144 specific monoclonal antibodies reacted with commensal gut bacteria of both human and mouse and showed limited neutralizing activities

To probe the potential antigens that might induce the P144 binding antibodies, we isolated 6 mAbs from two naïve SPF mice (one C57BL/6J mouse and one BALB/c mouse) with high levels of pre-existing S2 specific antibody responses. The results of microscale thermophoresis (MST) assays showed that the binding affinities with S2 protein were similar among the 6 mAbs (F5, 2.07 μM; H9, 0.98 μM; E10, 3.53 μM; G13, 2.10 μM; M3,1.46 μM; G18, 2.25 μM) (Supplementary Figure 5). Five of these mAbs recognized P144 solely (Supplementary Figure 6A), while one mAb (clone M3) bound with P144 and P103 simultaneously (Supplementary Figure 6B). Results of competitive ELISA showed that the minimal epitopes varied slightly among the five P144 specific mAbs, especially at the C-terminal of P144 (Supplementary Figure 6A). The neutralizing potentials of these isolated monoclonal antibodies were evaluated using a pseudo virus-based neutralization assay. Our results showed that these monoclonal antibodies exhibited limited neutralizing activity against 5 SARS-CoV-2 variants (Supplementary Figure 7), which might be partially explained by their relatively low affinities to S2 protein (Supplementary Figure 5).

To prove the cross-reactivities between S2 and commensal gut microbial antigens, whole cell lysates (WCL) of mouse and human commensal gut bacteria were prepared and used as antigens for WB assays, respectively. As shown in Figure 5A, specific bindings with the WCL of mixed fecal bacteria prepared from mice either with low levels of pre-existing antibodies (L) or with high levels of pre-existing antibodies (H) could be clearly visualized for each isolated mAbs. It was noteworthy that all mAbs except E10 strongly recognized a band around 180KD in the sample from mice with high pre-existing antibody responses. E10 predominantly recognized a band around 55KD in both samples, while stronger binding with the sample from mice with high pre-existing antibody responses could be visually observed (Figure 5A). Among the six mAbs, F5 showed the most diverse binding compacity. In addition to the band around 180KD, F5 bound with a band around 55KD (similar with E10) and a band between 40KD-55KD (Figure 5A). In comparison with the WB results of mouse samples, the recognized bands were less consistent across different human fecal bacteria samples (Figure 5B), presumably due to the individual to individual variation of gut microbiota composition. We found that a band around 70KD was recognized by most mAbs in 4 (lanes 1, 5, 6, 7) out of 7 samples and a band between 50KD-70KD was recognized by all mAbs in 3 (Lanes 2, 3, 7) out 7 samples. Our data showed that the recognition patterns towards each fecal bacteria sample were generally stable across different mAbs except E10 (Figure 5A and 5B). Then, we performed V(D)J gene sequencing for the 6 antibody clones. Our data showed that the same VH/VL gene combination (IGHV2-5*01, IGKV6-23*01) is used by 4 clones (F5, G18, H9 and M3) (Supplementary Table 1). Interestingly, among these 4 clones, F5 and M3 were isolated from a C57 mouse, while G18 and H9 were isolated from a BALB/c mouse, suggesting it might be a public antibody shared by different mice. G13 uses the same VH gene as the above 4 mAbs in combination with a different VL gene (IGLV2*02). The VH/VL gene usage (IGHV5-9*02, IGKV8-30*01) of E10 is completely different from other clones, which might explain its unique WB patterns shown in Figure 5A and 5B.

**Figure 5.**
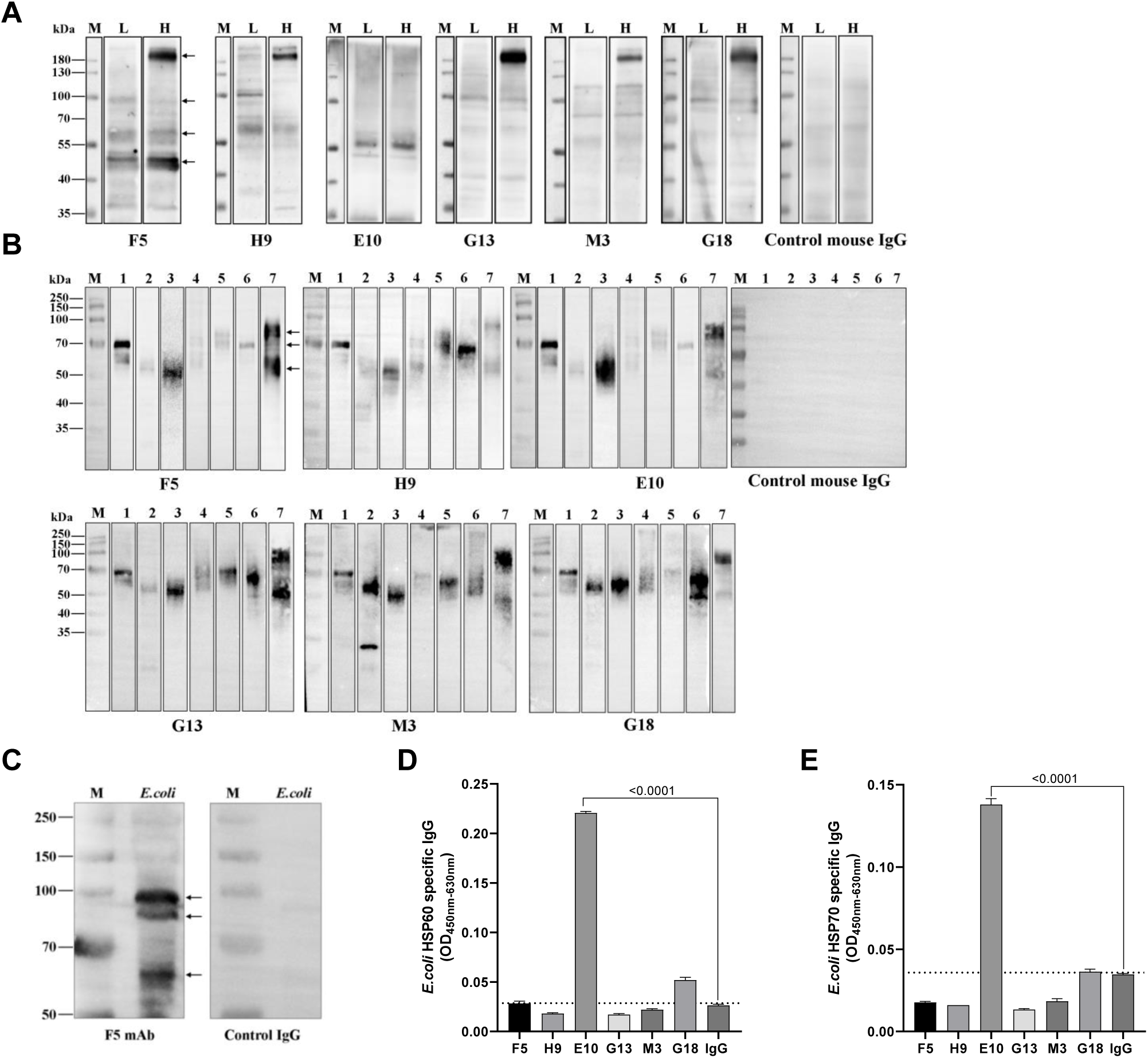
P144 binding mAbs isolated from naïve SPF mice reacted with commensal gut bacteria from both human and mouse. Reactivities between P144 binding mAbs and gut microbial antigens were detected using WB assays. A purified mouse IgG was used as the control. (**A**) WB assays of mouse fecal bacteria samples. L: mixed fecal bacteria samples collected from 3 mice with low levels of pre-existing S2 binding antibodies (OD_450nm-630nm_≤0.140, 1:100 diluted); H: mixed fecal bacteria samples collected from 3 mice with high levels of pre-existing S2 binding antibodies (OD_450nm-630nm_ ≥0.615, 1:100 diluted). (**B**) WB assays of fecal bacteria samples collected from 7 healthy individuals (Lanes 1-7). All the mAbs and the control mouse IgG were tested at the final concentration of 1μg/ml. Black arrows indicate the locations of protein bands selected for the mass spectrometry analysis. (**C**) Reactivity between a P144 specific mAb (clone F5) and *E. coli* was validated by WB. F5 or a purified mouse IgG was used as the first antibody at the concentration of 1 μg/ml. Black arrows point out the bands with MWs equal to *E. coli* proteins identified by LC-MS analysis. (**D**) Recognition of purified *E.coli* HSP60 protein by P144 reactive antibodies. (**E**) Recognition of purified *E.coli* recombinant HSP70 protein by P144 reactive antibodies. All the mAbs and control IgG were measured at a final concentration of 10 μg/ml. Dotted line represents the mean OD value of control IgG plus 2-fold SD (mean+2SD).

Proteins corresponding to specifically recognized bands were excised from Coomassie blue stained gels and analyzed by the mass spectrometry. For the mouse fecal bacteria samples, protein bands with molecular weights around 180KD, 100KD, 55KD-70KD and 40KD-55KD (indicated by arrows in Figure 5A, panel F5) were selected. For human fecal bacteria samples, protein bands with molecular weights around 50KD-70KD, 70KD and 70KD-100KD (indicated by arrows in Figure 5B, panel F5) were selected. The lists of proteins identified in mouse and human samples were shown in Table 2 and Table 3, respectively. Proteins with molecular weights corresponding approximately to the excised protein bands were identified for both human and mouse fecal bacteria samples. Of note, multiple proteins within the theoretical MW range of 58KD to 60KD were found to be identical between mouse and human samples, which included Fumarate hydratase class I (Accession# P14407), Formate-tetrahydrofolate ligase OS (Accession# Q189R2), Phosphoenolpyruvate carboxykinase (ATP) OS (Accession# C4ZBL1 and A6LFQ4) and 60 kDa chaperonin OS (Accession# A0Q2T1). To verify the cross-reactivity of the proteins detected by LC-MS, we selected *E. coli* (DH5α strain) as a representative target because *E. coli* derived Fumarate hydratase class I (Accession# P14407) were found in both human and mouse fecal bacteria samples. The result of WB assay showed that the P144 specific mAb (Clone F5) recognized multiple bands with MWs consistent with *E. coli* proteins identified by LC-MS (Figure 5C). Employing an in-house ELISA method, we further proved that one of the isolated mAbs (E10) bound specifically with purified microbial HSP60 and HSP70 proteins (Figure 5D and 5E), while it did not obviously bind with purified human HSP60 or HSP70 (Supplementary Figure 8).

**Table 1.**
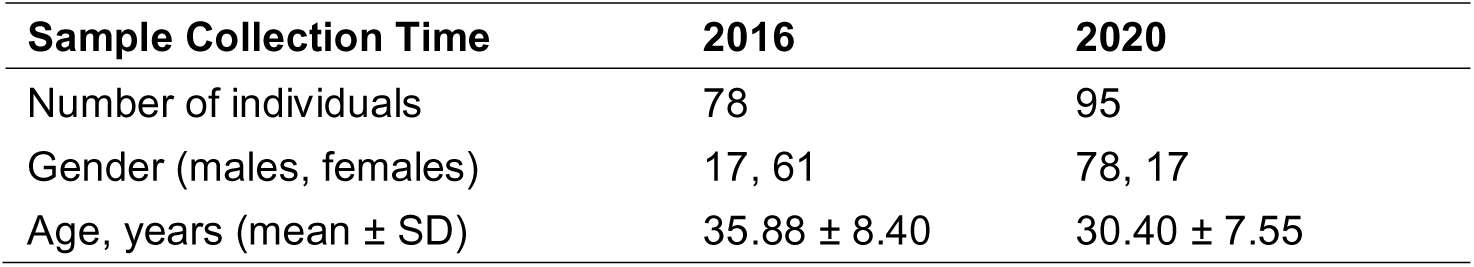
Demographics of SARS-CoV-2 unexposed healthy individuals

**Table 2.**
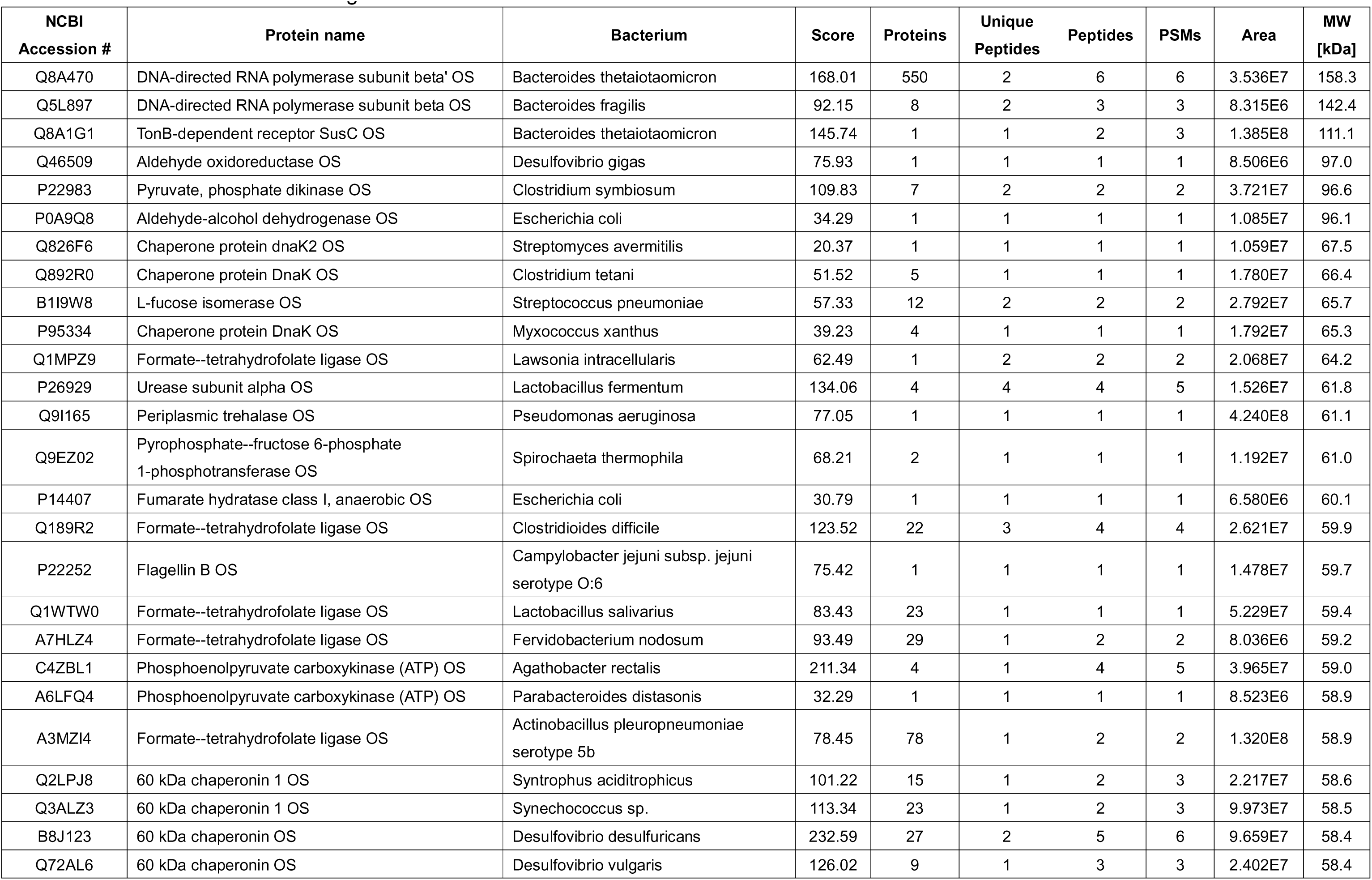
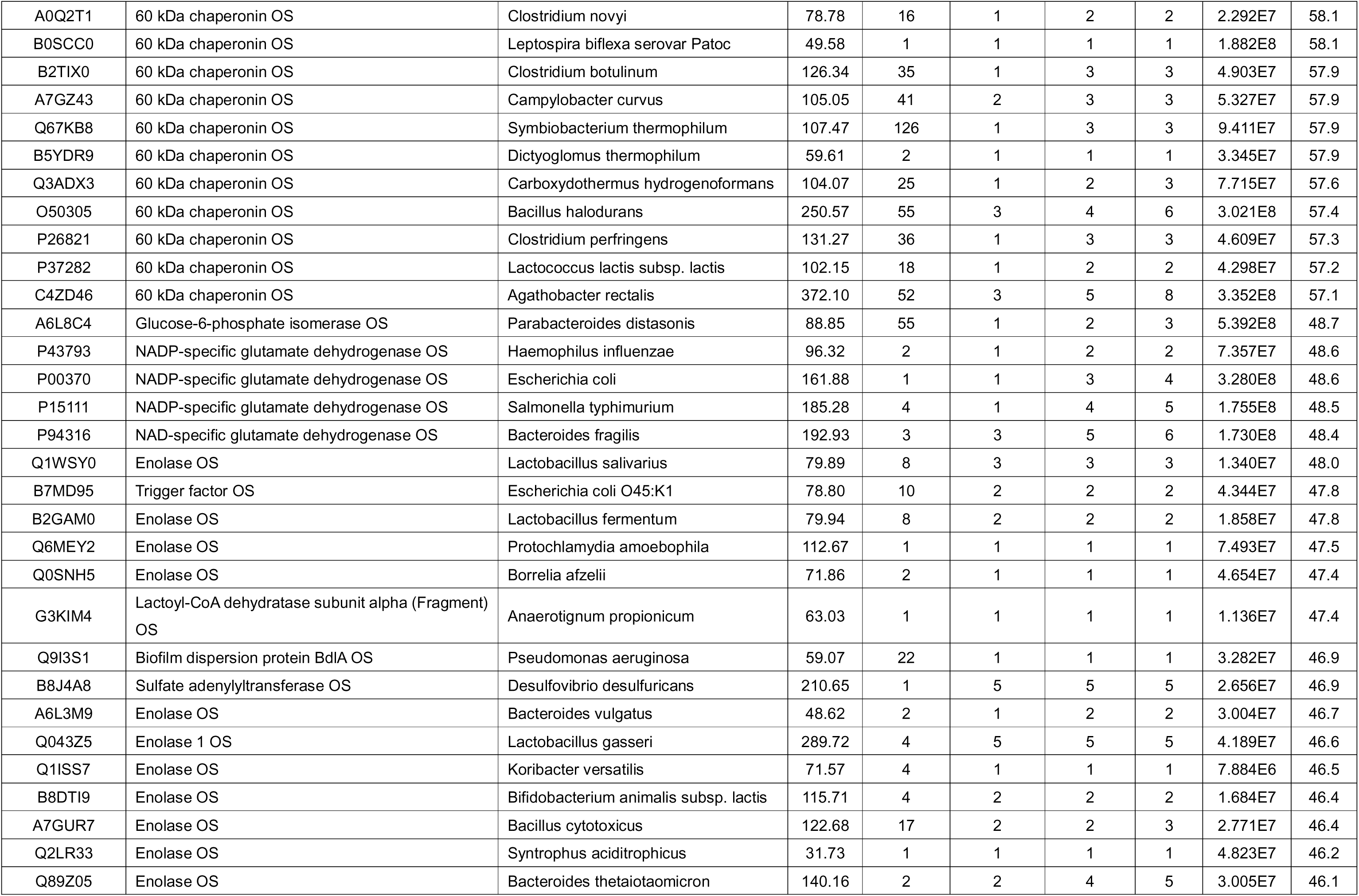
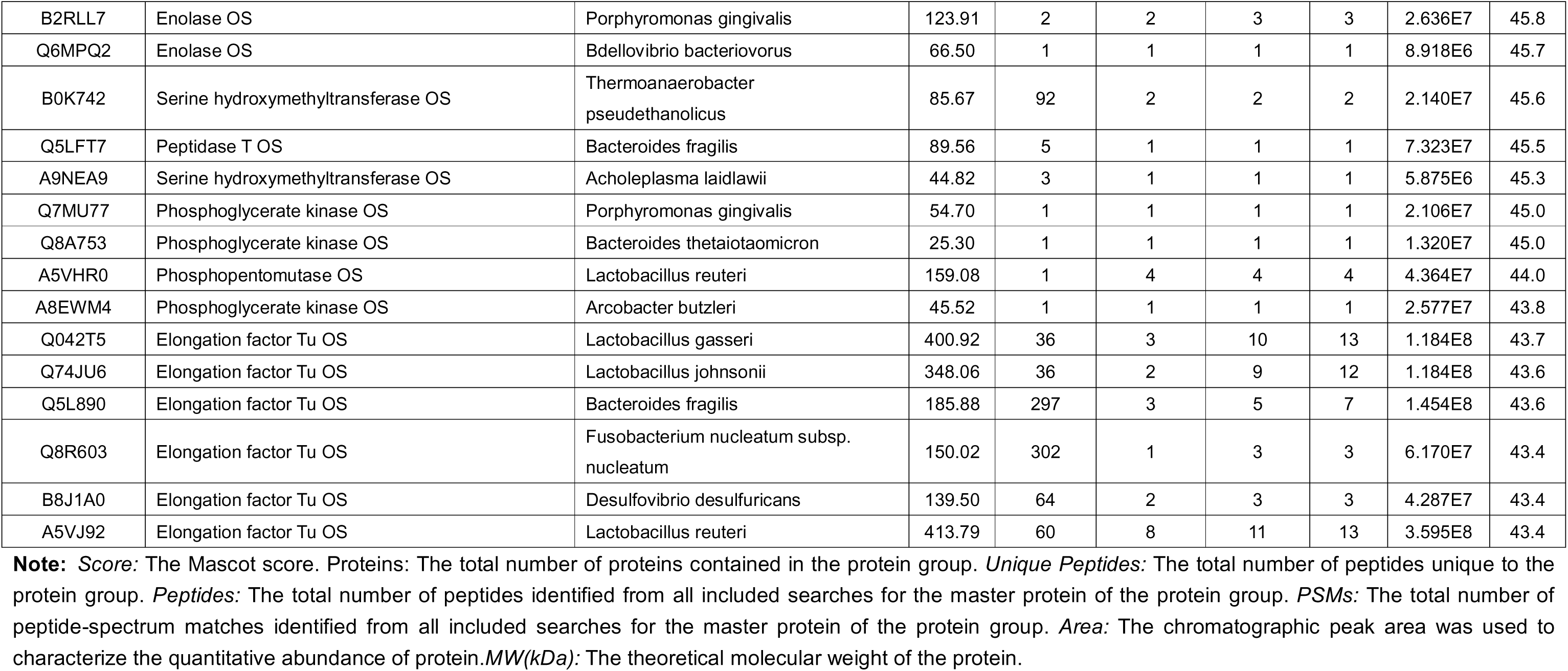
Potential cross-reactive antigens identified in mouse fecal bacteria

**Table 3.**
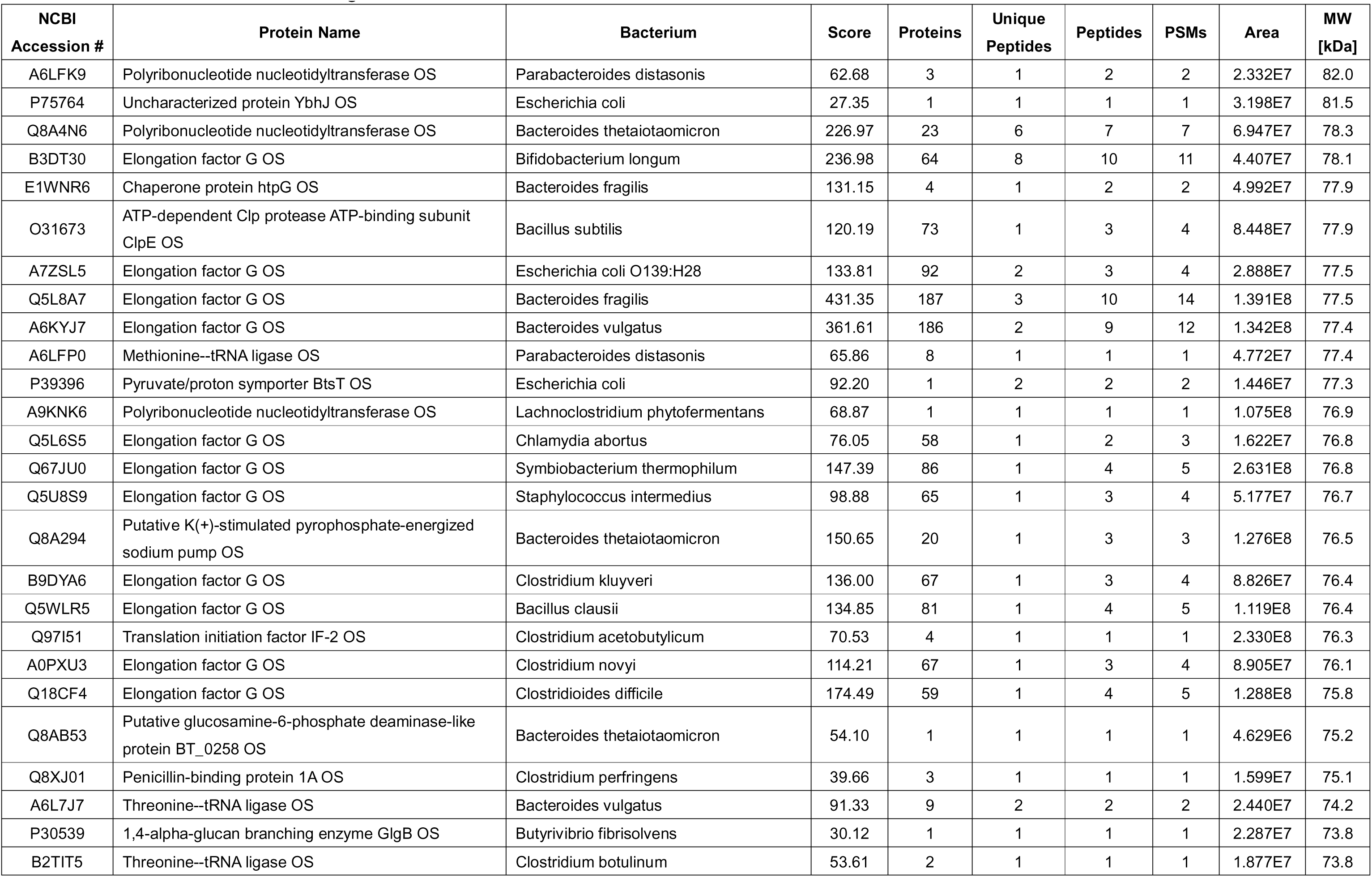
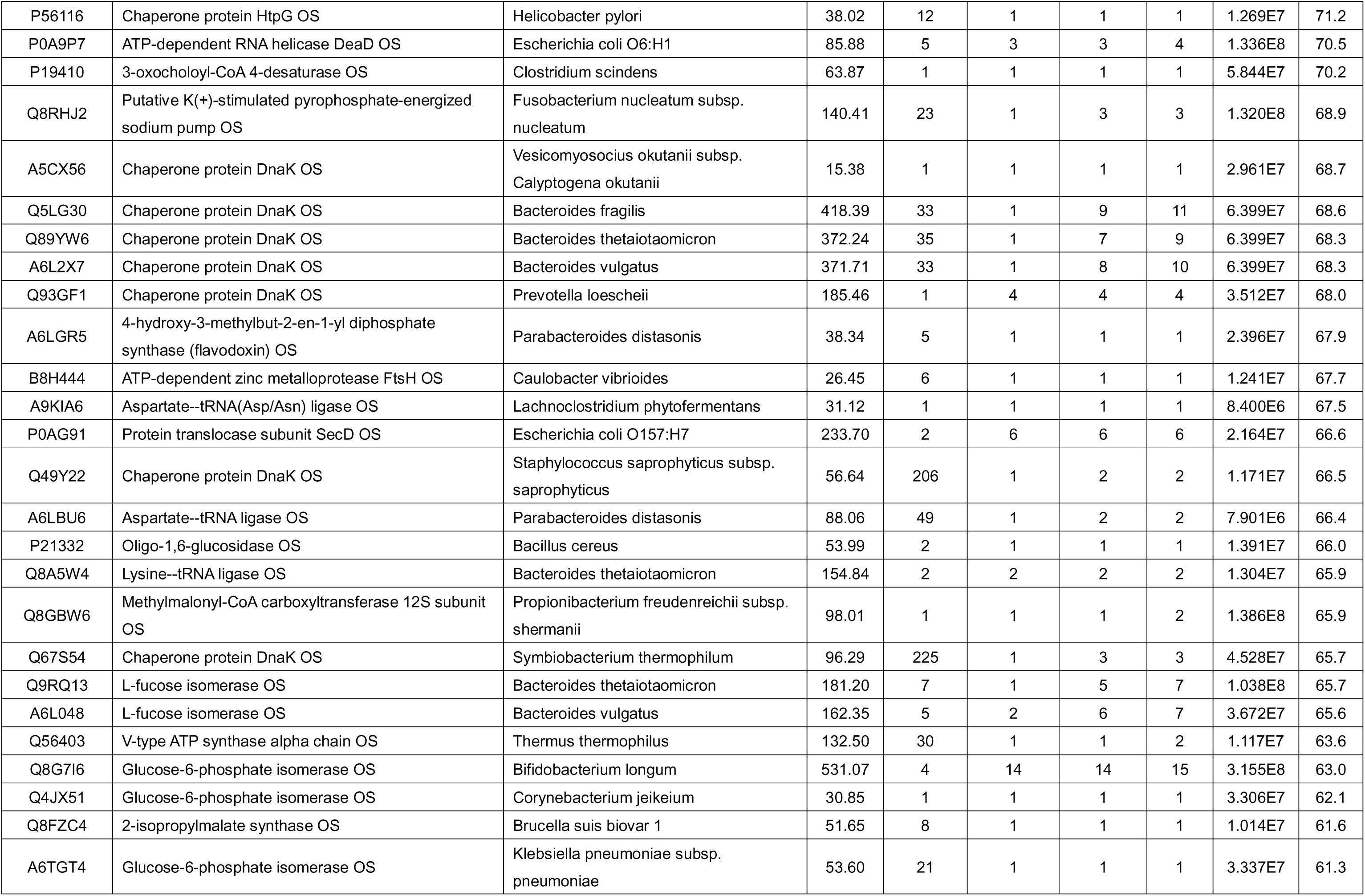
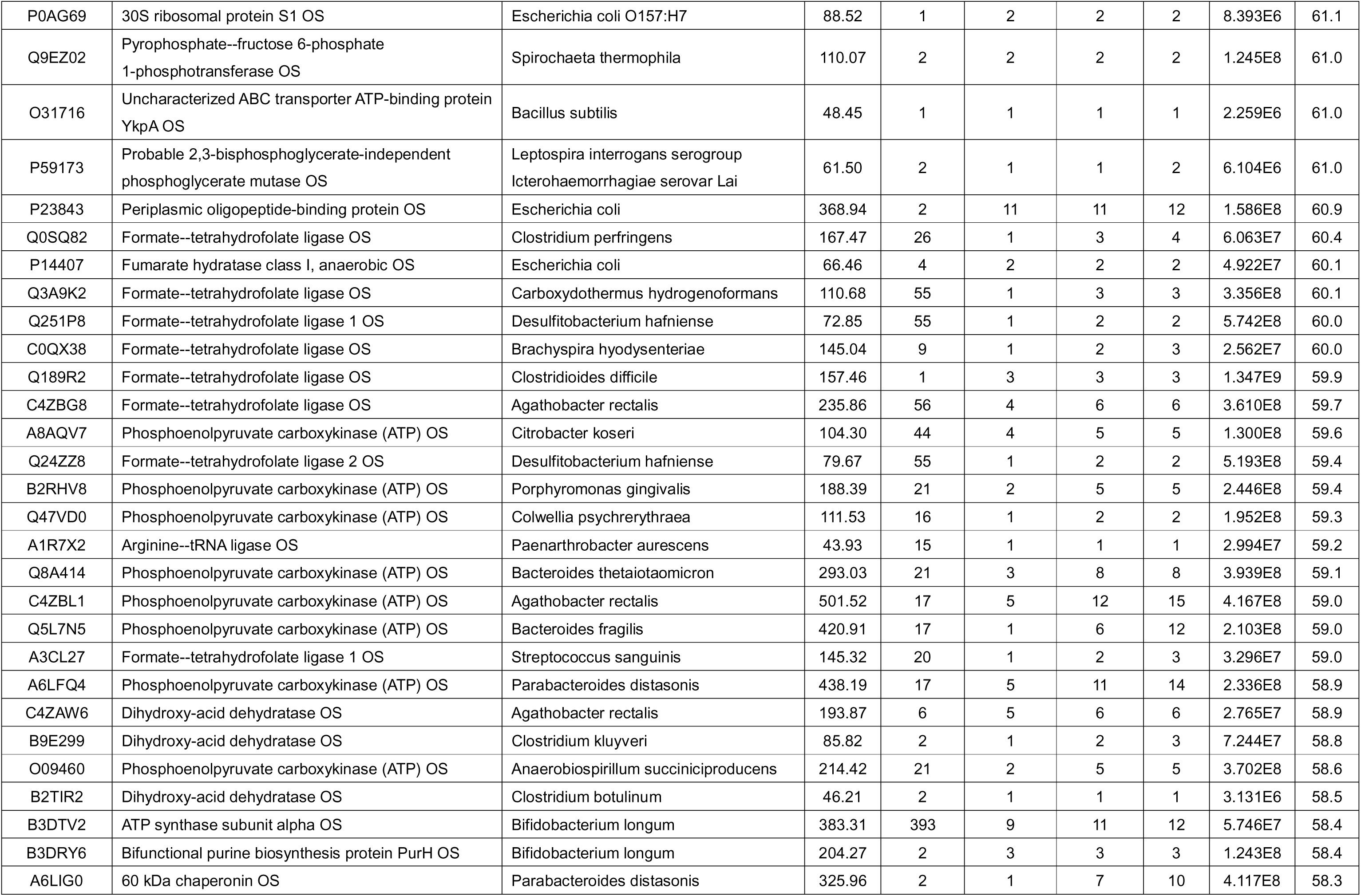
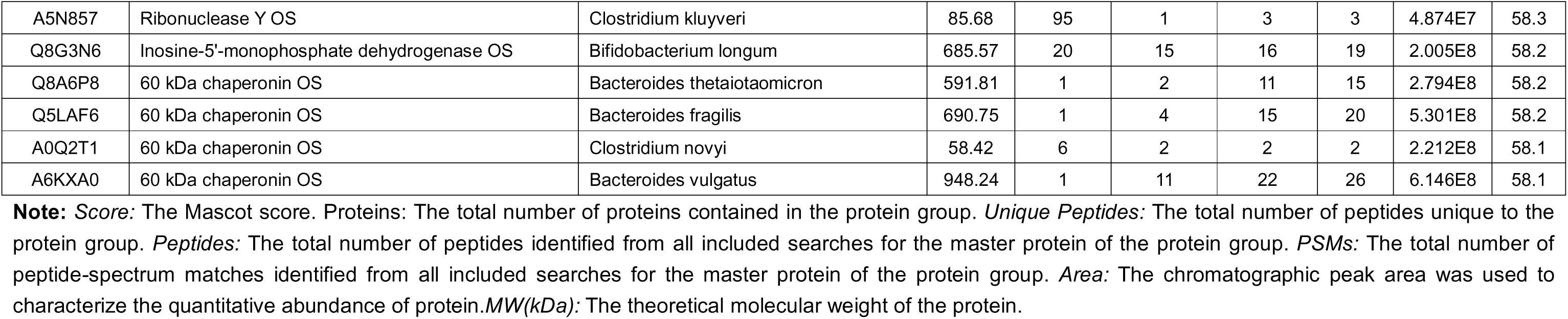
Potential cross-reactive antigens identified in human fecal bacteria

### Pre-existing S2 cross-reactive antibodies impacted specific immunities induced by a candidate COVID-19 DNA vaccine in mice

Pre-existing antibodies has been shown to be able to shape the recall immune responses upon influenza infection and vaccination (6). And the concern about how the pre-existing immunities may influence the effect of a SARS-CoV-2 vaccine has also attracted lots of attention (33). To investigate the impact of the pre-existing P144 antibodies on the immunogenicity of a candidate DNA vaccine, 18 BALB/c mice were divided into 3 groups according to their levels of pre-existing S2 binding antibodies and immunized with a DNA vaccine encoding the full length of SARS-CoV-2 S protein (Figure 6A and 6B). Our data showed that mice with high levels of pre-existing S2 binding antibodies mounted significantly higher S2 binding antibody responses after vaccination compared to mice with low or moderate levels of pre-existing S2 binding antibodies (Figure 6C). The average level of P144 specific antibody responses was also stronger in mice with high levels of pre-existing S2 binding antibodies than that of mice with low pre-existing S2 binding antibody responses (Figure 6D and 6E). By comparison, both the S1 binding antibody and the neutralizing antibody titers did not significantly differ among all groups, despite that mice with moderate or high levels of pre-existing antibodies tended to mount higher average titer of S1 binding antibodies (Figure 6F and 6G). Mice without vaccination showed neither obvious S1 binding antibody response nor neutralizing activity (Data not shown). We further investigated the influence of pre-existing antibodies on humoral immune responses in mouse respiratory tract after vaccination. And our data showed that the levels of S1 specific IgG in BALF were similar among the three groups after DNA vaccination (Figure 7A), while the average level of S2 specific IgG in BALF from mice with high pre-existing S2 binding antibodies was significantly higher than those from mice with low pre-existing antibodies (Figure 7B). S protein specific IgA response did not increase significantly after vaccination as compared with unvaccinated group (Figure 7C and 7D).

**Figure 6.**
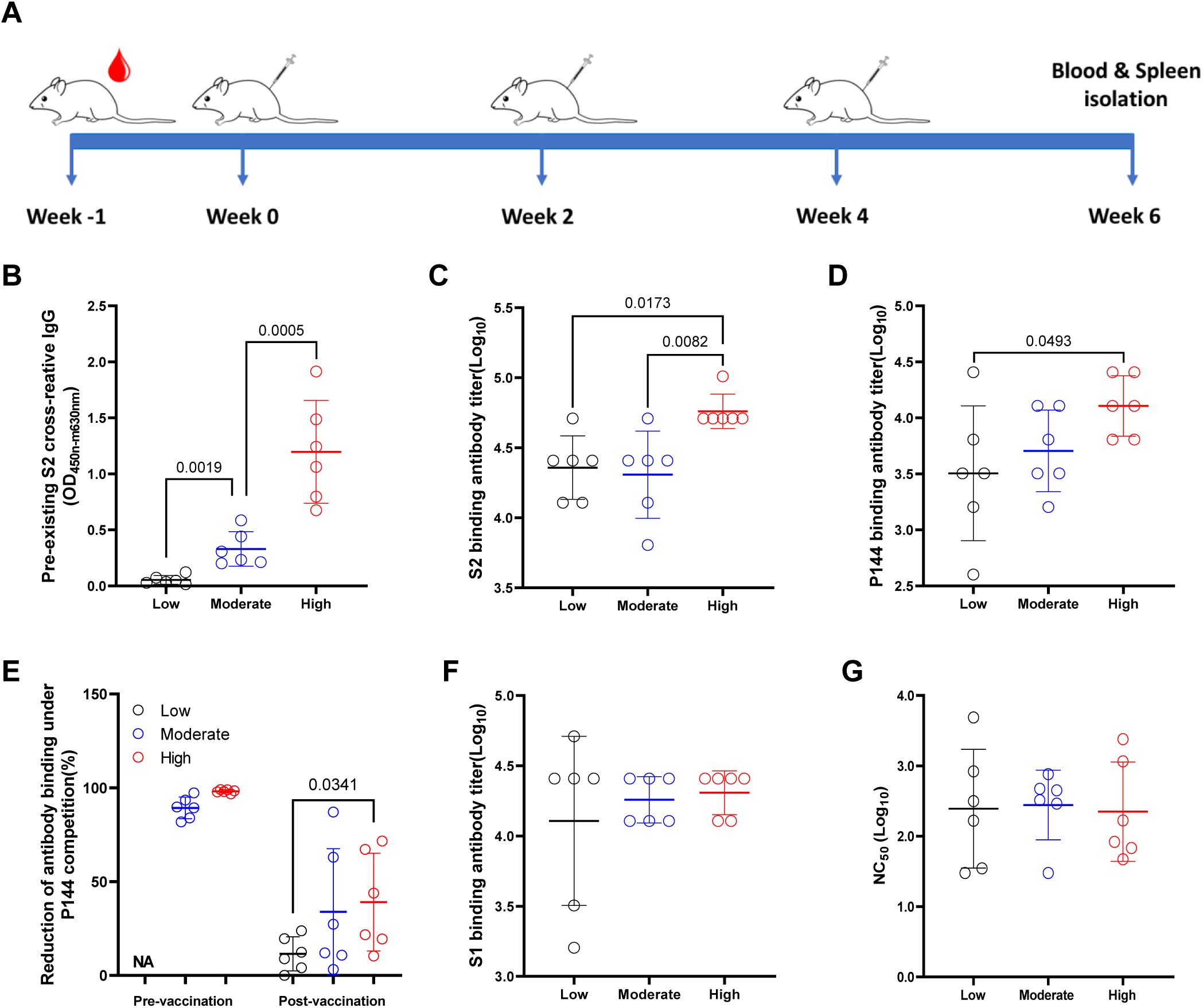
Impacts of pre-existing antibodies on the humoral immune responses elicited by a DNA vaccine encoding SARS-CoV-2 S protein. (**A**) Schematic illustration of the experiment schedule. 50μg of the DNA vaccine was injected intra muscularly into each mice at week 0, week 2 and week 4, respectively. Two weeks after the final vaccination, the mice was euthanized for the measurements of specific immune responses. (**B**) Peripheral blood was collected before immunization and levels of pre-existing S2 specific antibodies were compared among groups. (**C**) Endpoint IgG titers against S2 were compared at 2 weeks post the last immunization. (**D**) Comparisons of P144 specific IgG titers as measured using BSA-P144 conjugate as the coating antigen. (**E**) Comparisons of P144 specific binding antibody levels as determined using a method of competitive ELISA. (**F**) Endpoint IgG titers against S1 measured at 2 weeks post the final vaccination. (**G**) Neutralizing antibody titers against SARS-CoV-2 D614G pseudo-virus in serum of mice at 2 weeks post the final immunization. Data were shown as mean±SD, n=6. Statistical analyses were performed by the method of one-way ANOVA.

**Figure 7.**
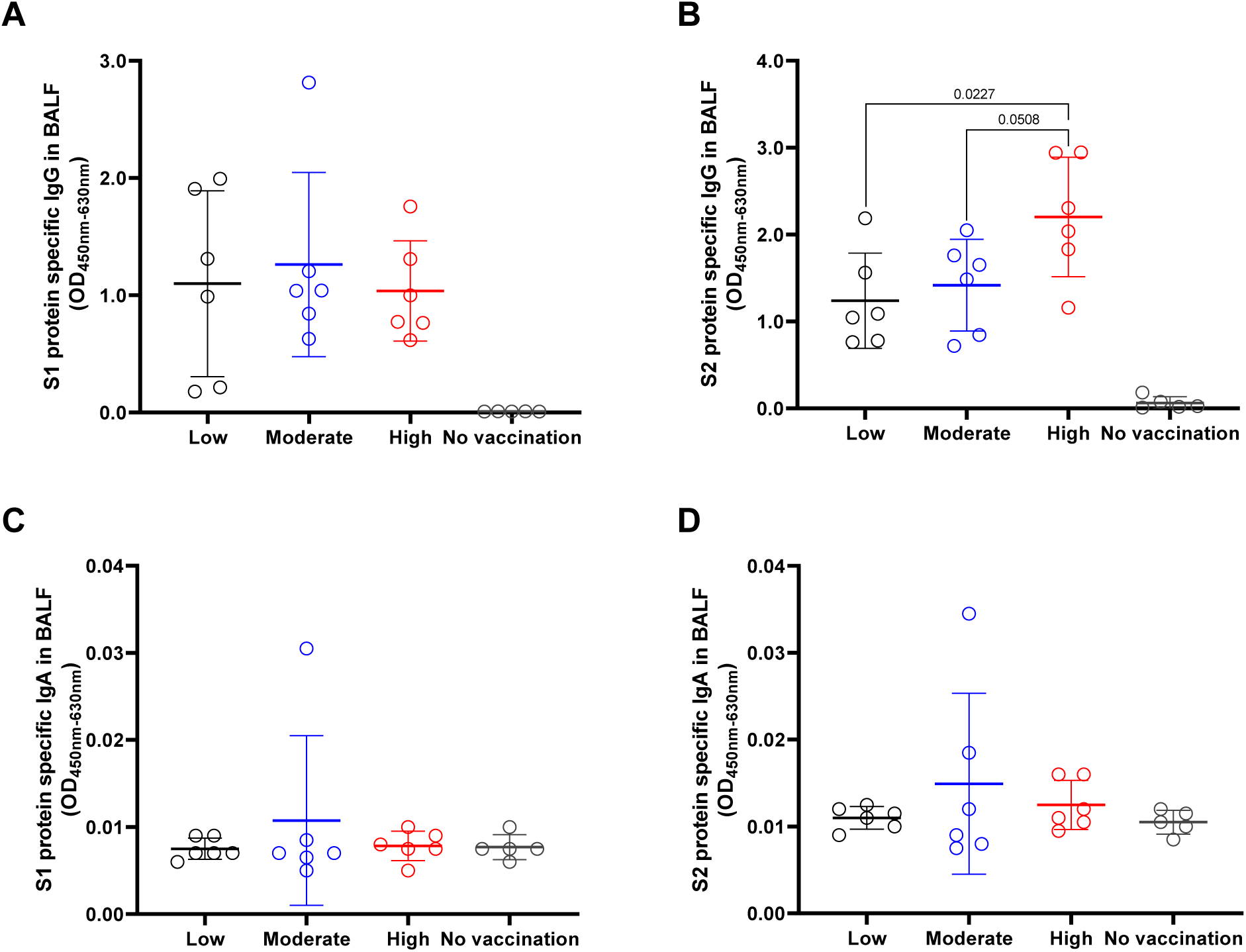
The impacts of pre-existing antibody on the levels of specific IgG and IgA in BALF after vaccination. BALF was collected from each mouse after euthanasia. Specific IgG (**A** and **B**) and IgA (**C** and **D**) against S1 or S2 were detected using in-house ELISA methods. All the BALF samples were adjusted to the initial total protein concentration of 51.9μg/ml. Data are shown as mean±SD, n=6. Statistical analyses were performed by the method of one-way ANOVA.

As the pre-existing antibodies in naïve SPF mice predominantly recognized P144 (Figure 2C and Supplementary Figure 1), we delineated the impact of pre-existing antibodies on the recognition of this epitope after vaccination. Our results showed that the minimum epitope recognition pattern by the sera of mice with high levels of pre-existing antibodies remained unchanged after vaccination (Figure 8). Whereas the minimum epitope recognized by the sera of mice with moderate and low levels of pre-existing antibodies altered at either the N-terminal or both terminals of P144 (Figure 8).

**Figure 8.**
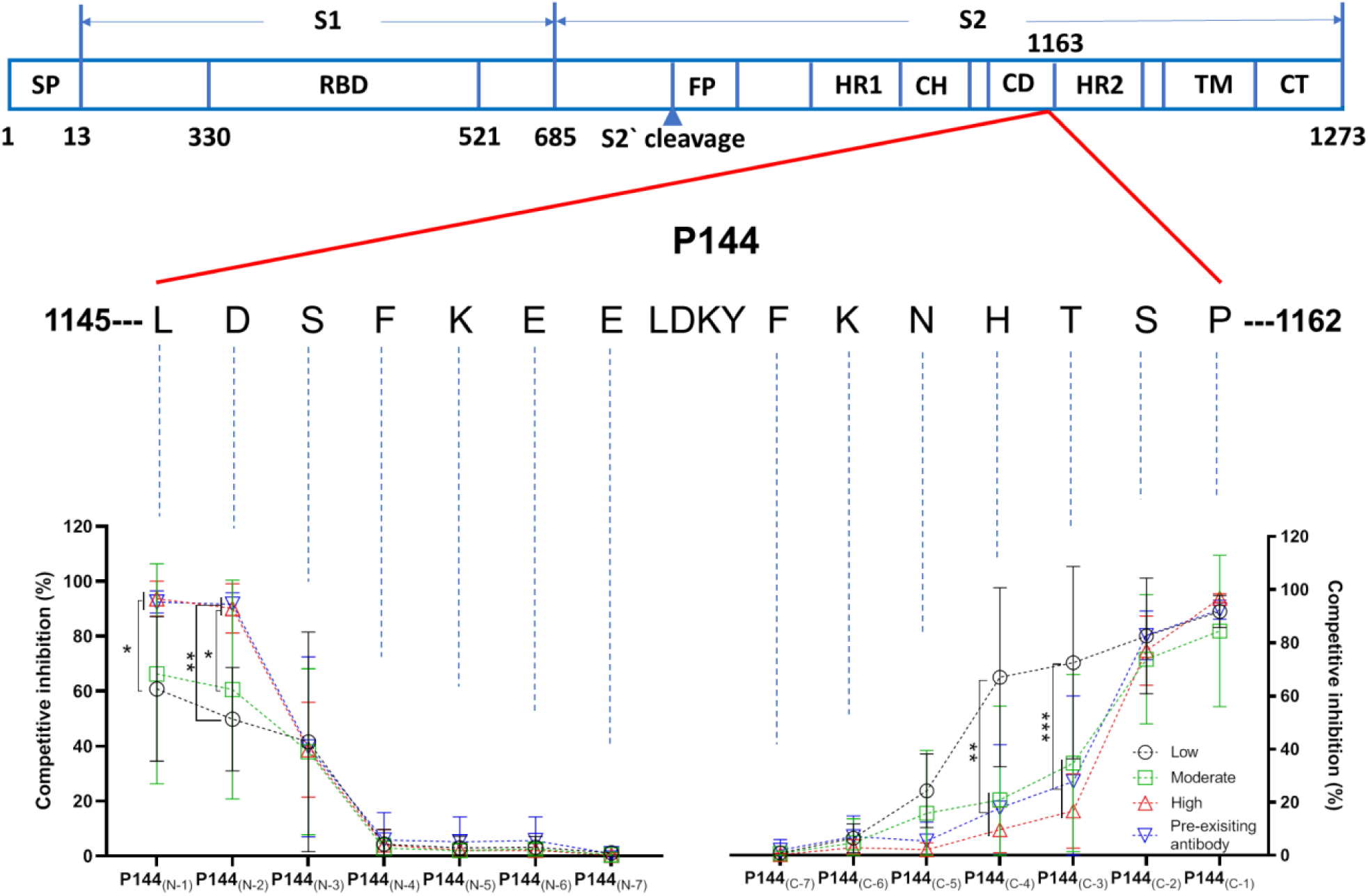
The impact of pre-existing antibody on the recognition of the minimum epitope of P144 after vaccination. The minimal epitope recognized by mouse sera after vaccination was analyzed using a method of competitive ELISA. Purified BSA-P144 conjugate was used as the coating antigen and truncated peptides derived from P144 were used as the competitors. The decreases of competitive inhibition reflected the necessity of each amino acid for the epitope recognition. Statistical analyses were performed by the method of two-tailed t-test (*, P< 0.05, **, P< 0.01, ***, P< 0.001).

In addition to antibody measurement, we compared S protein specific T cell responses among the three groups as well (Figure 9A). The results showed that the candidate DNA vaccine elicited robust S protein specific T cell responses in all groups (Figure 9). Although no statistical significance was reached, interesting trends were observed: First, mice with high levels of pre-existing S2 binding antibodies tended to mount relatively stronger S1 and S2 specific IFN-γ+ T cell responses (Figure 9B, 9C and 9D); second, as measured by the releases of IL-6, IL-2 and TNF-α, mice with high levels of pre-existing antibodies tended to mount stronger T cell responses against both S1 and S2 (Figure 9C and 9D). Mice without vaccination showed no S protein specific T cells responses (Data not shown). The major findings of this part of the study were validated by a repeated experiment (Supplementary Figure 9).

**Figure 9.**
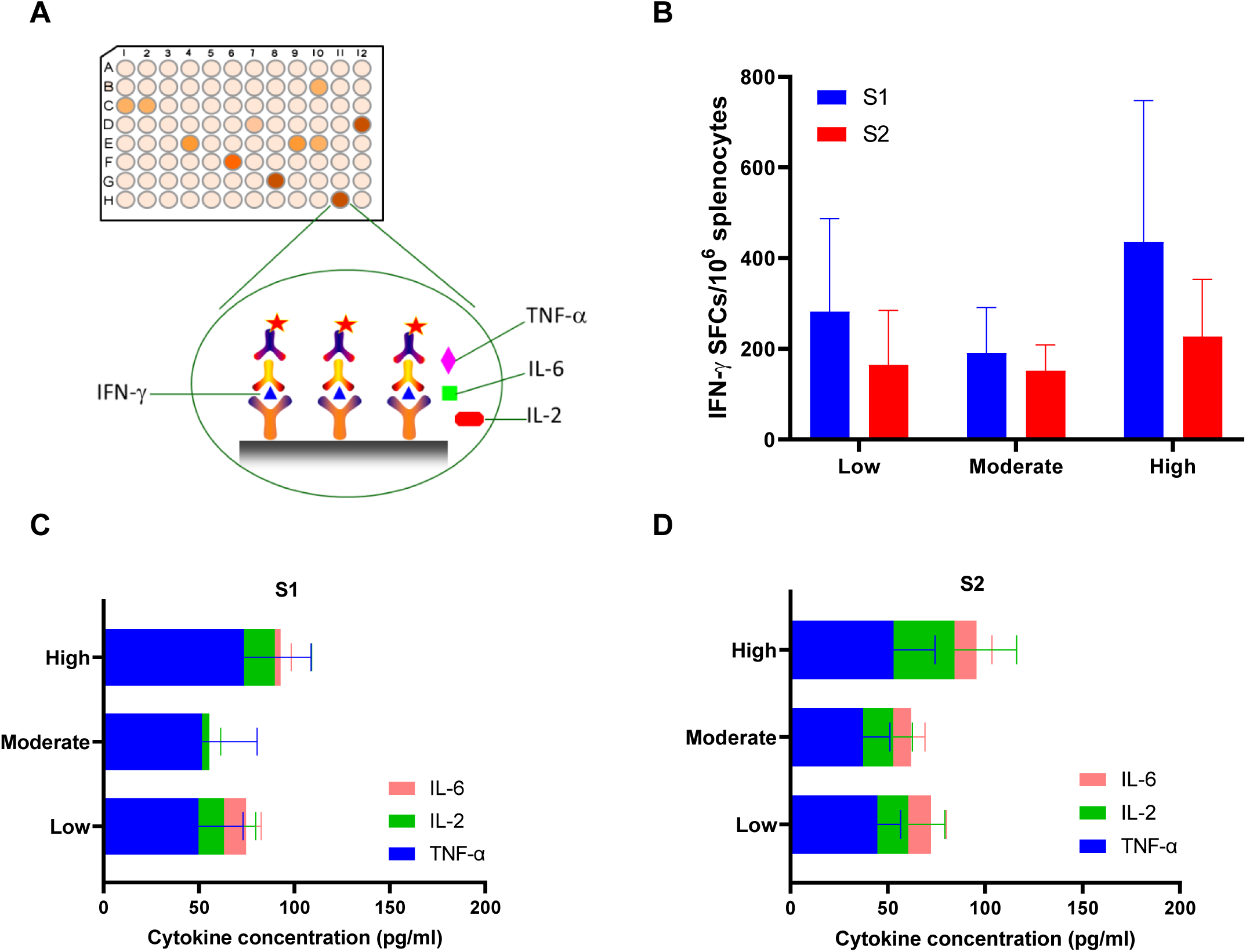
The impacts of pre-existing antibody on the cellular immune responses after vaccination. (**A**) The diagram of the method used for cellular immune responses evaluation. (**B**) S1 and S2 specific IFN-γ responses were compared among groups of mice with different levels of pre-existing S2 reactive antibodies. Additionally, S1 (**C**) and S2 (**D**) specific releases of IL-2, IL-6 and TNF-α as measured using the method of multiplex cytokine bead assay were also compared among different groups. Data were shown as mean± SD, n=6. SFCs, spot forming cells.

### Pre-existing S2 cross-reactive antibodies correlated with RBD binding antibody responses after two-dose inactivated SARS-CoV-2 vaccination

To investigate how the pre-existing cross-reactive antibodies may influence the COVID-19 vaccine induced immunity, peripheral blood samples were collected from 28 healthy individuals who received two doses of an inactivated SARS-CoV-2 vaccine (Figure 10A). Correlation analyses showed that both the OD values (Figure 10B and 10C) and the titers (Supplementary Table 2) of pre-existing S2 reactive antibodies were significantly associated with RBD binding antibody titers at 14 days after immunization. Additionally, although not statistically significant, the pre-existing P144 binding antibody levels tended to correlate positively with neutralizing antibody responses after vaccination (P=0.0946) (Figure 10D).

**Figure 10.**
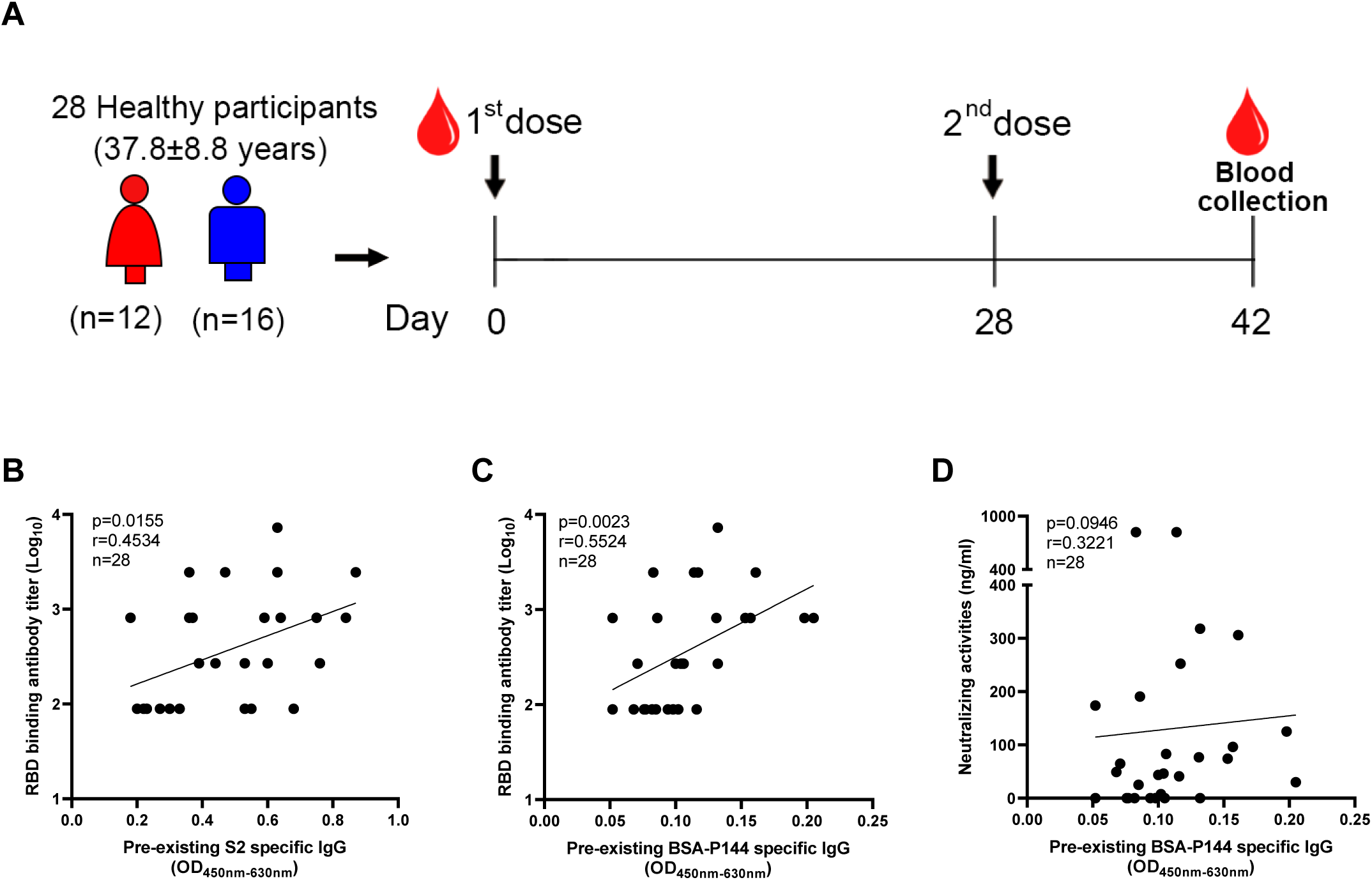
Levels of pre-existing S2 reactive antibodies correlated with RBD binding antibody responses elicited by an inactivated SARS-CoV-2 vaccine in human. **(A)** Peripheral blood samples were collected from 28 healthy vaccines who received two doses of an inactivated SARS-CoV-2 vaccine at baseline and 14 days post the 2^nd^ dose, respectively. The RBD binding antibody titers were measured by ELISA. The neutralizing antibody responses were quantified by a commercialized surrogate virus neutralization test (sVNT) (Suzhou Sym-Bio Life Science Co., Ltd). **(B and C**) Correlations between RBD binding antibody titers and levels of pre-existing S2 or P144 specific IgG. **(D)** Correlation between neutralizing antibody concentrations and pre-existing P144 binding antibody levels. Statistical analyses were performed using the method of Spearman’s correlation. The experiment was repeated twice.

### Impact of SARS-CoV-2 S DNA vaccination on the compositions of mouse gut microbiota

To explore whether SARS-CoV-2 S DNA vaccination can affect the composition of gut bacteria, mouse stool samples were collected before and after vaccination and analyzed by 16s rDNA sequencing (Supplementary Figure 10A). The results showed that P144 specific antibody increased significantly after DNA vaccination (Supplementary Figure 10B). The PCA analysis showed that the bacterial genera composition of vaccinated mice clearly clustered together (Supplementary Figure 10C). More specifically, at the genus level, abundances of candidatus saccharimonas and muribaculaceae increased significantly after DNA vaccination, while no significant increase was observed in PBS group (Supplementary Figure 10D).

## Discussion

The origins of pre-existing cross-reactive immunities against SARS-CoV-2 have been investigated vigorously since the outbreak of the pandemic (39). Accumulating data suggest that cross-reactive T cells (33, 40–43) in SARS-CoV-2 unexposed human might be induced by previous infections of other hCoVs. While the origins of pre-existing cross-reactive antibodies could not be completely explained by previous infections of other coronaviruses, as recent studies revealed that the magnitude of antibody responses to SARS-CoV-2 S protein in the sera of patients with COVID-19 was not related to HCoVs’ S titers (44) and immunization with coronaviruses OC43 did not induce significant SARS-CoV-2 S protein cross-reactive antibodies in mice. Moreover, it has also been observed that SARS-CoV-2 S protein specific binding antibody responses were weak in SARS-CoV-2 unexposed individuals with obvious binding antibody responses against S proteins of common cold hCoVs (45, 46).

To track the potential origins of the pre-existed cross-reactive antibodies targeting SARS-CoV-2 spike protein, in this study, we first screened the cross-reactive antibody responses in SARS-CoV-2 unexposed human plasma collected in 2020 and 2016, respectively. In both cohorts, we found that the magnitudes of S2 binding antibodies were significantly higher than those of S1 binding antibodies. This finding is consistent with previous studies showing that pre-existing S2 cross-reactive antibody responses are stronger than S1 cross-reactive antibody responses in SARS-CoV-2 unexposed individuals (26, 44, 47, 48). Since S2 cross-reactive antibody responses have also been observed in unexposed animals (44), we continued to screen the cross-reactive antibody responses in two strains of naïve SPF mice. Our data showed that the OD values of S2 cross-reactive antibodies were significantly higher than those of S1 cross-reactive antibodies in naïve BALB/c and C57BL/6 mice. Detections of mouse sera collected from another two independent SPF animal facilities confirmed this finding (Data not shown). We also tried to detect the SARS-CoV-2 S protein specific T cells responses in mice with high pre-existing S2 cross-reactive antibodies using the IFN-γ ELISPOT assay, which showed that there was no pre-existing cross-reactive T cell response among these mice (Data not shown).

To facilitate the search of potential antigens that induced the cross-reactive antibodies, we identified a dominant antibody epitope (P144) through a method of competitive ELISA based linear antibody epitope mapping. P144 is located within the connector domain of S2 (aa1147-aa1160, directly N-terminal of the HR2 region). The same epitope has been predicted (30) and detected in both SARS-CoV-2 unexposed and infected individuals by multiple previous studies (22, 23, 25, 26, 32). In this study, we detected P144 specific antibody responses in plasma samples of healthy individuals collected in both 2020 and 2016. More interestingly, we found that the pre-existing S2 cross-reactive antibodies in naïve SPF mice were predominantly against this epitope. Of note, although the aa sequence of P144 is highly conserved among SARS-CoV-2 variants and SARS-CoV, its similarity with four seasonal hCoVs is relatively low. It has also been shown that this epitope was more frequently recognized than its homologous peptides from common cold hCoVs by antibodies of COVID-19 negative individuals (23). These evidence collectively implied that the pre-existing S2 specific antibodies might not be necessarily elicited by previous common cold coronavirus infections.

To unveil the origin of the pre-existing S2 binding antibodies in mice, we first measured S2 specific B cells by B cell ELISPOT and flowcytometry and found that the frequency of S2 specific B cells was significantly higher in mesenteric LN than in spleen, suggesting that the gastrointestinal tract might be the primary site where the cross-reactive B cells were activated. As gut bacteria can promote B cell diversification and stimulate antibody production in both T-dependent and -independent ways (49), we speculated that exposure to certain gut microbial antigens might account for the presence of the cross-reactive antibodies. To prove this hypothesis, we interrogated the relationship between the levels of pre-existing S2 reactive antibodies and the compositions of mouse gut microbiota. Our results showed that mice with different gut microbiota composition had different levels of pre-existing S2 cross-reactive antibodies, and vice versa (Supplementary Figure 4). Moreover, we found that transplantation of fecal bacteria isolated from SPF mice could induce S2 reactive antibodies in mice bred in a sterile isolation pack. The above evidence suggested that the S2 reactive antibodies could be induced by exposure to certain commensal gut bacteria. Nonetheless, the gut microbiota might not be the only factor that can affect the generation of the cross-reactive antibodies, because we observed that mice housed in the same cage could harbor different levels of S2 reactive antibodies (Data not shown).

We are not able to define the bacterial strains contributing to the induction of P144 reactive antibodies in the current study, because it is technically hard to get pure cultures for each potential strain. Instead, we tried to identify potential microbial antigens that may induce the cross-reactive antibodies. To do so, we isolated six P144 specific monoclonal antibodies from a naïve BALB/c mouse and a naïve C57BL/6 mouse, respectively. All the six mAbs were confirmed to be able to bind with P144 and showed weak neutralizing compacities against five SARS-CoV-2 variants. Leveraging these mAbs, we detected the cross-reactive antigens in mouse and human fecal microbiota through WB assays. Compared with a control mouse IgG, specific bands were observed for each mAb, which proved the antibody cross-reactivities between SARS-CoV-2 and commensal gut bacteria. The strongly recognized protein bands were further analyzed by LC-MS. Our data showed that cross-reactive antigens derived from bacteroides and parabacteroides were frequently identified in fecal bacteria samples of both human and mouse, which was consistent with our metagenomic sequencing data showing that the abundance of bacteroides and parabacteroides was significantly higher in the commensal gut bacteria of mice with high pre-existing S2 binding antibody levels. More intriguingly, five cross-reactive microbial antigens were identified in mouse and human fecal samples simultaneously, implying that the S2 cross-reactive antibodies might naturally occur in different species of mammals.

According to our LC-MS results, few proteins of *E.coli* and HSP60/HSP70 proteins of multiple bacterial strains can be recognized by P144 reactive antibodies. These results were verified by experiments showing that P144 reactive mAbs could bind with the lysate of an E.coli strain (DH5α) and purified HSP60/HSP70 proteins. However, due to the limited availabilities of pure cultures of commensal gut bacteria and their protein derivatives, we were not able to verify all the potential reactive strains in this study. Alternatively, we tried to refine our LC-MS results through sequence similarity analysis. We found that P144 shared varied identities with LC-MS captured proteins (40%-70%, length≥8 aa residues) (Data not shown). The similarities are not high enough to support confident identifications of potential cross-reactive epitopes based on our current data. To clarify this issue, we plan to expand experimental screening through collaboration in future.

In parallel with tracking the initial antigens that induced the S2 cross-reactive antibodies, we investigated the impact of pre-existing antibodies on the immunogenicity of a candidate DNA vaccine as well. According to previous reports, pre-existing cross-reactive antibodies may influence the effects of different vaccines differentially (6, 50). In this study, we found that the pre-existing cross-reactive antibodies shaped the vaccine-induced immune responses in both mouse and human. Mice with high levels of pre-existing antibodies mounted stronger S2 binding antibodies in both peripheral blood and bronchial lavage after vaccination. More interestingly, we found that the pre-existing antibody levels correlated positively with post-vaccination RBD binding antibody titers in human. These findings proved that the pre-existing S2 binding antibodies could facilitate the generation of vaccine induced antibody responses. Through epitope mapping, we observed that the pre-existing antibodies strongly restricted the minimal epitope recognition in mice with high levels of pre-existing antibodies, which suggested that the imprint effect of pre-existing cross-reactive antibodies on vaccine induced antibody responses was primarily epitope specific. In addition to antibody response, we also found that the high levels of pre-existed S2 binding antibodies tended to improve specific T cell responses induced by SARS-CoV-2 S DNA vaccine. Since we did not perform the live virus challenge, it is still not clear how the pre-existing S2 cross-reactive antibodies will impact vaccine efficacy *in vivo*. Nonetheless, as both our results and a recently published study suggested that antibodies targeting P144 epitope could neutralize SARS-CoV-2 (51), we speculate that the pre-existing P144 cross-reactive antibodies may have protective effect. Besides, in this part of the study, we tried but found it may not be appropriate to test the influences of pre-existing antibodies on vaccine elicited immunities through passive antibody transfer, because: First, passive antibody transfer cannot generate S2 reactive memory B cells in recipient mice, which was observed in the mesenteric lymph nodes and spleens of mice with pre-existing S2 reactive antibodies (Supplementary Figure 3). Second, we found that in vivo antibody level dropped dramatically within 48 hours after tail vein injection and it was technically difficult to maintain relatively stable in vivo antibody level for a long term.

A deep understanding of pre-existing cross-reactive antibodies against SARS-CoV-2 will enable better therapeutic, diagnostic and vaccine strategies. In this study, we provided evidence showing that antibodies targeting a conserved linear epitope on S2 cross-reacted with gut microbial antigens from both human and mouse, manifesting that some of the pre-existing cross-reactive antibodies might be induced by exposure to certain commensal gut bacteria. We proved that the pre-existing S2 cross-reactive antibodies did not impair the immunogenicity of a candidate DNA vaccine in a mouse model, while the SARS-CoV-2 S DNA vaccination could significantly change the composition of mouse gut microbiota. Further investigations into the functions of P144 cross-reactive antibodies may assist in delineating the role S2 specific antibody upon SARS-CoV-2 infection and elucidating the mechanisms underlying the gastrointestinal symptom caused by COVID-19 (52–54).

## Materials and methods

### Ethics statement

All experiments and methods were performed in accordance with relevant guidelines and regulations. Experiments using mice and samples of healthy human were approved by the Research Ethics Review Committee of the Shanghai Public Health Clinical Center Affiliated to Fudan University.

### Plasma samples of healthy human

Two batches of plasma samples were collected from healthy individuals at the health screening clinic of Shanghai Public Health Clinical Center. A concurrent batch was collected in December 2020. All the 95 individuals enrolled in this batch reported no epidemiological link with confirmed COVID-19 patients and were confirmed to be free from any chronic or acute disease. Viral RNA tests confirmed that all individuals in this batch were free from SARS-CoV-2 infection. In addition, a historical batch of 78 plasma samples from healthy individual cohort (collected in 2016) were also measured for their cross reactivities with SARS-CoV-2 S protein. As the local prevalence in Shanghai was extremely low during previous SARS-CoV-1 epidemic, we did not do the serum screening for previous SARS-CoV-1 infection. Instead, we collected the information regarding previous SARS-CoV-1 infection status through either questionnaire survey (for the 2020 cohort) or telephone follow-up (for the 2016 cohort). There is no self-reported previous SARS-CoV-1 infection among the two cohorts. Demographical information about these two cohorts was described in Table 1.

### Detection of SARS-CoV-2 S1 and S2 specific binding antibodies

In-house enzyme-linked immunosorbent assays (ELISA) were developed to measure SARS-CoV-2 S1 and S2 specific binding antibodies. High-binding 96-well EIA plates (Cat# 9018, Corning, USA) were coated with purified SARS-CoV-2 S1 (Cat# 40591-V08H, Sino Biological, China), S2 proteins (Cat# 40590-V08B, Sino Biological, China), recombinant *E.coli* HSP60 (Cat#HSP-004, Prospec, Jsrael), *E.coli* HSP70 (Cat#HSP-006, Prospec, Jsrael), human HSP60 (Cat#HSP-016, Prospec, Jsrael) or human HSP70 (Cat#HSP-170, Prospec, Jsrael) at a final concentration of 1µg/ml in carbonate/bi-carbonate coating buffer (30mM NaHCO_3_,10mM Na_2_CO_3_, pH 9.6). Subsequently, the plates were blocked with 1× PBS containing 5% milk for 1 hour at 37°C. Next, 100μl of diluted human plasma, mouse serum or mAbs was added to each well. After 1-hour incubation at 37°C, the plates were washed with 1× PBS containing 0.05% Tween20 for 5 times. Then, 100 µl of a HRP labeled rabbit anti-human IgG antibody (Cat# ab6759, Abcam, UK) or goat anti-mouse IgG antibody (Cat# 115-035-003, Jackson Immuno Research, USA) diluted in 1× PBS containing 5% milk were added to each well and incubated for 1 hour at 37°C. After a second round of wash, 100μl of TMB substrate reagent (Cat# MG882, MESGEN, China) was added to each well. 15 minutes later, the color development was stopped by adding 100μl of 1M H_2_SO_4_ to each well and the values of optical density at OD450_nm_ and OD630_nm_ were measured using 800 TS microplate reader (Cat# 800TS, Biotek, USA).

### Competitive ELISA

According to the reference sequence of SARS-CoV-2 (Genebank accession number: NC_045512), peptides (18-mer overlapping by 11 residues, purities >95%) encompass the full length of S protein were synthesized by GL Biochem (Shanghai, China). The experiment procedure was generally similar with the afore mentioned in-house ELISA assays, except that the diluted mouse serum or human plasma were incubated with synthesized peptides (5μg/ml) or a non-relevant peptide (OVA_323-339_) for 1 hour at room temperature before adding into the coated EIA plates.

### FACS analysis of S2 specific B cells in mice

Spleen and mesenteric lymph nodes were isolated from naïve SPF mice and single-cell suspensions were freshly prepared. After counting, 1 × 10^6^ single cells were resuspended in 100µl R10 (RPMI1640 containing 10% fatal bovine serum) and incubated with biotinylated S2 protein (Cat# 40590-V08B-B, Sino Biological, China) for 30 minutes at room temperature. After incubation, the cells were washed twice with 500µl R10. Then, the cells were incubated with the mixture of PE-anti-mouse CD19 (Cat# 152408, Biolegend, USA, 1µl/test), BV510-anti-mouse CD45 (Cat# 103137, Biolegend, USA, 1.25µl/test) and Streptavidin-IF647 (Cat# 46006, AAT Bioquest, USA, 0.2µl/test) at room temperature for 30 minutes. After washing, the stained cells were resuspended in 200µl 1×PBS and analyzed using a BD LSRFortessa™ Flow Cytometer. The data were analyzed using the FlowJo software (BD Biosciences, USA).

### Preparation of P144 specific monoclonal antibodies

Monoclonal antibodies against P144 were prepared from one naïve BALB/c mouse and one naïve C57BL/6 mouse respectively using the hybridoma technique. Briefly, freshly isolated splenocytes were mixed and fused with SP2/0 cells at a ratio of 1:10. Hybridoma cell clones secreting P144 specific antibodies were screened by ELISA and monoclonal hybridoma cells were selected by multiple rounds of limited dilution. Selected clones of hybridoma cells were injected intraperitoneally into BALB/c×ICR hybrid mice. About 1-2 weeks later, peritoneal fluid was collected, and monoclonal IgG was purified using Protein A resin. The purities of monoclonal antibodies were verified using SDS-PAGE and the antibody concentrations were determined using a BCA kit (Cat# P0012, Beyotime Biotechnology, China).

### V(D)J gene sequencing of P144 reactive monoclonal antibodies

The V(D)J genes of P144 reactive monoclonal antibodies were sequenced by AZENTA life science. Briefly, total RNA was extracted from hybridoma cells using Trizol reagent (Cat# R4801-02, Invitrogen, USA). 5’ RACE was performed with SMARTer RACE cDNA Amplification Kit (Cat# 634923, Clontech, USA), total RNA input was 500-2000ng. V(D)J genes of heavy and light chains were amplified by PCR. The PCR products were purified through gel extraction using a QIAquick Gel Extraction Kit (Cat# 28704, Qiagen, USA). NGS libraries were constructed by using VAHTS Universal DNA Library Prep Kit for Illumina (Cat# ND607, Vazyme, China). The qualified libraries were sequenced on the Illumina Miseq 2×300 platform (Illumina, San Diego, CA, USA). Raw fastq files were first subject to quality assessment. Adapters and bases with poor quality scores (Q value lower than 20) were removed using Trimmomatic (v0.36) to generate clean data (trimmed data). Pandaseq (2.10) was used to merge pair-end read. Merged sequences were processed by IgBLAST software to identify the V(D)J sequences. The reference sequences were obtained in IMGT database (IMGT, https://www.imgt.org/).

### Isolation of gut commensal bacteria and preparation of whole cell lysate (WCL)

About 2g of each fecal sample was suspended with 15ml sterile 1×PBS and vortexed thoroughly to obtain uniform mixtures. After centrifugation at 200×g for 5 min, the supernatants were collected, and the sediments were discarded. This process was repeated twice. Next, all the supernatant samples were centrifuged twice at 9000×g for 5 min and the supernatants were discarded. The precipitated bacteria pellets were resuspended in 500µl of 1×PBS (containing 1mM PMSF) and disrupted with an ultrasonic cell crusher (the probe-type sonicator, Model JY92-II; Ningbo Scientz Biotechnology Co., Ltd, China). After sonication, the samples were centrifuged at 10000rpm for 30 minutes to remove the cellular debris.

### Western blotting

WCL containing 10μg of total protein was separated by SDS-PAGE (10% acrylamide gels) and then transferred onto a PVDF membrane (Cat# IPVH00010, Millipore, USA) or stained with Coomassie brilliant blue. After blocking with 5% skim milk for 2h, the membrane was incubated with a P144 specific monoclonal antibody or a control mouse IgG at a concentration of 1 μg/ml. After washing, the membrane was incubated with HRP conjugated rabbit anti-human IgG antibody (Cat# ab6759, Abcam, UK) or HRP conjugated goat anti-mouse IgG antibody (Cat # 115-035-003,Jackson Immuno Research, USA) diluted 1:5000 in TBST (Tris-buffered saline, pH 8.0, 0.05% Tween 20) containing 5% skim milk. After wash, the bands were developed with an ultra-sensitive ECL substrate (Cat# K-12045-D10, Advansta, USA). The area corresponding to the specific WB bands were excised from the gel stained with Coomassie blue and analyzed using the mass spectrometry.

### Mass spectrometry analysis

The FASP digestion was adapted for the following procedures in Microcon PL-10 filters (Cat#MRCPRT010, Merck, USA). After three-time buffer displacement with 8 M Urea (Cat#U111898, Aladdin, China) and 100 mM Tris-HCl, pH 8.5, proteins were reduced by 10 mM DTT (Cat#646563, Sigma Aldrich, USA) at 37 °C for 30 min and followed by alkylation with 30 mM iodoacetamide at 25°C for 45 min in dark. Digestion was carried out with trypsin (enzyme/protein as 1:50) (Cat#T9201, Sigma Aldrich, USA) at 37°C for 12 h after a wash with 20% ACN (Cat#34851, Sigma Aldrich, USA) and three-time buffer displacement with digestion buffer (30 mM Tris-HCl, pH 8.0). After digestion, the solution was filtrated out and the filter was washed twice with 15% ACN, and all filtrates were pooled and vacuum-dried to reach a final concentration to 1 mg/ml. LC-MS analysis was performed using a nanoflow EASYnLC 1200 system (Thermo Fisher Scientific, Odense, Denmark) coupled to an Orbitrap Fusion Lumos mass spectrometer (Thermo Fisher Scientific, Bremen, Germany). A one-column system was adopted for all analyses. Samples were analyzed on a home-made C18 analytical column (75 µm i.d. × 25 cm, ReproSil-Pur 120 C18-AQ, 1.9 µm (Dr. Maisch GmbH, Germany). The mobile phases consisted of Solution A (0.1% formic acid (Cat#695076, Sigma Aldrich, USA) and Solution B (0.1% formic acid in 80% ACN). The derivatized peptides were eluted using the following gradients: 2–5% B in 2 min, 5–35% B in 100 min, 35–44% B in 6 min, 44–100% B in 3 min, 100% B for 10 min, at a flow rate of 200 nl/min. Data-dependent analysis was employed in MS analysis: The time between master scan was 3s, and fragmented in HCD mode, normalized collision energy was 30.

### Construction and preparation of a candidate DNA vaccine encoding SARS-CoV-2 full length S protein

The full-length *s* gene sequence of the reference SARS-CoV-2 strain was optimized according to the preference of human codon usage and synthesized by GENEWIZ life science company (Suchow, China). The codon optimized spike gene was subcloned into a eukaryotic expression vector (pJW4303, kindly gifted by Dr. Shan Lu’s Laboratory at the University of Massachusetts) (55, 56). And the sequence of inserted gene was verified by Sanger sequencing (Sangon Biotech Co., Ltd., Shanghai, China). An EndoFree Plasmid Purification Kit (Cat#12391, Qiagen, Hilden, USA) was used to prepare the recombinant plasmid for mouse vaccination.

### Mouse vaccination

Peripheral blood samples were collected from female adult mice and pre-existing S2 binding antibodies were measured using the previously described in-house ELISA method. According to their pre-existing S2 binding antibody levels (at 1:100 dilution of serum), the mice were divided into three groups: low (0.015 < OD_450nm-630nm_ ≤ 0.130, n=6), moderate (0.130< OD_450nm-630nm_ ≤ 0.750, n=6) and high (OD_450nm-630nm_ > 0.750, n=6). All mice were immunized intramuscularly with the candidate S protein DNA vaccine (50μg/mouse) for three times at an interval of 2 weeks. Three weeks post the third vaccination, the mice were euthanized. Peripheral blood, bronchial lavage and spleen were collected for assays of S protein specific immune responses.

### Metagenomic and 16s rDNA sequencing of mouse gut microbiota

Metagenomic DNA and 16s rDNA were sequenced by SHANGHAI BIOCHIP CO., LTD. For metagenomic DNA sequencing, bacterial DNA was extracted from fecal samples using a TIANamp Stool DNA Kit (Cat#DP328, TIANGEN, China). Then, a total amount of 1μg DNA per sample was used as input material for the DNA sample preparations. Metagenomic sequencing libraries were generated using NEBNext® Ultra™ DNA Library Prep Kit for Illumina (Cat#E7103, NEB, USA) following manufacturer’s recommendations. Constructed libraries were analyzed for size distribution by Agilent 2100 Bioanalyzer and quantified using real-time PCR. The clustering of the index-coded samples was performed on a cBot Cluster Generation System. After cluster generation, the library preparations were sequenced on an Illumina Novaseq 6000 platform and paired-end reads were generated.

For 16s rDNA sequencing, fragments of 16s rDNA were amplified with specific barcoded primers 338F (5’-CCTAYGGGRBGCASCAG-3’) and 806R (5’-GGACTACNNGGGTATCTAAT-3’). After purification of the PCR product, sequencing libraries were generated using NEBNext® Ultra™ DNA Library Prep Kit for Illumina (Cat#E7654, NEB, USA) following manufacturer’s recommendations. After quality evaluation, the library was sequenced on an Illumina Novaseq 6000 platform and 250bp paired-end reads were generated.

### SARS-CoV-2 pseudo-virus neutralization assay

VSV-backboned SARS-CoV-2 pseudo-viruses were prepared according to a reported method (57). The neutralization assay was conducted by following the previously described procedure (57, 58). Briefly, 100μl of serially diluted mice sera were added into 96-well cell culture plates. Then, 50μl of pseudo-viruses with a titer of 13000 TCID_50_/ml were added into each well and the plates were incubated at 37°C for 1 hour. Next, Vero cells were added into each well (2×10^4^ cells/well) and the plates were incubated at 37°C in a humidified incubator with 5% CO_2_. 24 hours later, luminescence detection reagent (Bright-Glo™ Luciferase Assay System, Promega, USA) was added to each well following the manufacturer’s instruction. The luminescence was measured using a luminescence microplate reader (GloMax® Navigator Microplate Luminometer, Promega, USA) within 5 minutes. The Reed-Muench method was used to calculate the virus neutralization titer. Antibody neutralization titers were presented as 50% maximal inhibitory concentration (IC_50_).

### Detections of S protein specific cellular immune responses

SARS-CoV-2 S protein specific IFN-γ releases were measured using the method of enzyme-linked immunosorbent spot (ELISPOT) assays (Cat# 551083, BD Bioscience, USA) according to a previously described procedure (59). Briefly, the 96-well ELISPOT plates were coated with purified anti-mouse IFN-γ monoclonal antibody overnight at 4°C. Then, the plates were blocked and 2 × 10^5^ fresh splenocytes were added into each well and incubated with peptide pools for 20 hours at 37°C in a humidified incubator with 5% CO_2_. The final concentration for each peptide was 1μg/ml. After incubation, detecting antibody and Avidin-HRP were added sequentially. Finally, the plates were developed using the BD™ ELISPOT AEC Substrate Set (Cat#551951, BD Bioscience, USA) according to the manufacturer’s manual. Spots representing IFN-γ producing cells were enumerated using an automated ELISPOT plate reader (ChampSpot III Elispot Reader, Saizhi, Beijing, China). At the same time, the supernatants in the wells of ELISPOT plates were also collected for detecting secreted cytokines using a multiplexed cytokine beads array kit (Cat#741054, Biolegend, USA).

### Statistical analysis

All statistical analyses were performed using GraphPad Prism 8 (GraphPad Software, Inc., La Jolla, CA, USA). Comparisons between two groups were conducted by the method of *t*-test. Comparisons among three or more group were done using one-way ANOVA. *P*<0.05 was considered as statistically significant.

## Data Availability

All data generated or analyzed during this study are included in this article.

## Acknowledgements

We thank Miss Zhangyufan He from Huashan Hospital, Fudan University, for her kind help with the language polishing. This work was funded by the National Natural Science Foundation of China (Grant No. 81971559, 82041010, 81971900, 31872744), National Science and Technology Major Project (Grant No. 2018ZX10731301-004, 2018ZX10302302-002 and 2018ZX10301-404-002-003) and the Science and Technology Commission of Shanghai Municipality (Grant No. 20411950400).

## Author contributions

Y.M.W., Y.X., C.Q., Z.Q.Z. and W.H.Z. designed the study. L.Q.J., S.F.W., Y.M.W., X.X.T., Y.F.Z., J.W., X.Y.W., and J.W. conducted the experiments. Y.M.W., L.Q.J. and S.F.W. analyzed the data and drafted the manuscript. Y.M.W., Y.X., C.Q., Z.Q.Z., W.H.Z. and F.F. revised the manuscript. D.M.Y. and W.H.W. provided intellectual inputs in tackling technical challenges in tracking the potential cross-reacting antigens.

## Data availability

The data of metagenomic analysis of gut microbiota has been deposited to the NCBI Sequence Read Archive (SRA) database with the accession number PRJNA747837.

