## Supplementary Figures and Tables for "Pre-existing antibodies targeting a linear epitope on SARS-CoV-2 S2 cross-reacted with commensal gut bacteria and shaped vaccine induced immunity"

### Supplementary Figures and Figure legends

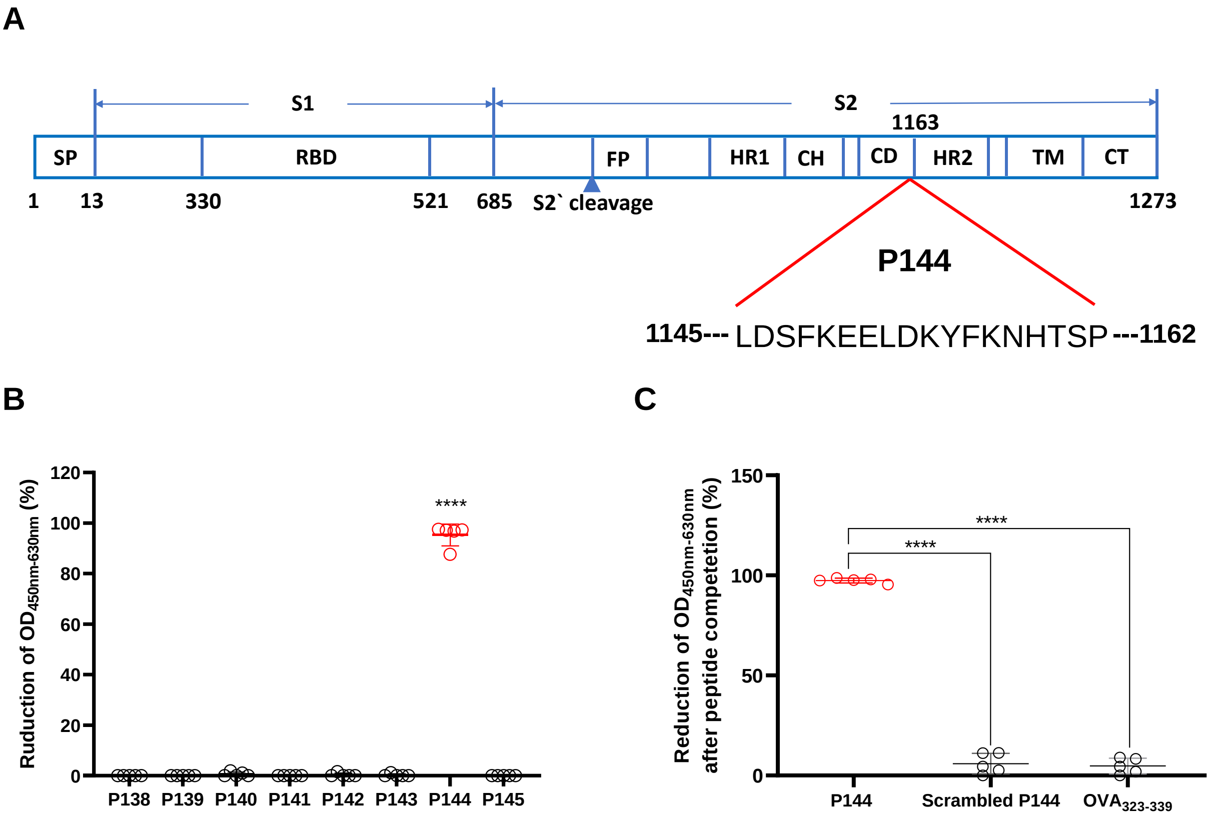


**Supplementary Figure 1 A dominant linear epitope recognized by the pre-existing antibodies locates on the connector domain of S2.** (**A**) Illustration of the location of P144 on the full length of SARS-CoV-2 spike protein. Synthesized peptides (18-mer, overlapped by 11amino acids) spanning the full length of S2 were divided into 9 peptide pools, each contained 8 peptides. To identify the potential antibody binding epitopes, we first performed competitive ELISA assays using the peptide pools as competitors (Data not shown). The pool showing significant inhibition was further delineated by testing the inhibiting efficiencies for each individual peptide. (**B**) A linear antibody epitope was identified via the method of competitive ELISA assay. (**C**) The specificity of P144 mediated inhibition was verified using a scrambled P144 (N`-LKHSKFDLNYKETSDEPF-C`) and a non-relevant peptide ($OVA_{323\text{-}339}$) as controls. The data were shown as mean±SD, n=5. Inhibition efficiencies among groups were compared using the method of One-way ANOVA. ****, p<0.0001. SP, signal peptide; RBD, receptor-binding domain; FP, fusion peptide; HR, heptad repeat; CH, central helix; CD, connector domain; TM, transmembrane domain; CT, cytoplasmic tail.

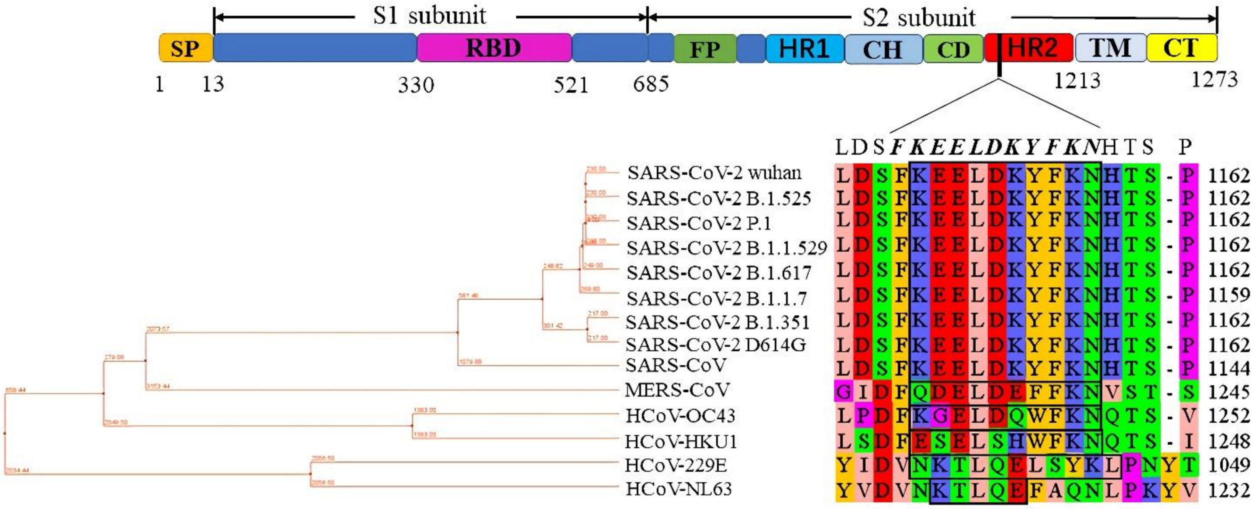


**Supplementary Figure 2 Phylogenetic analysis of the spike protein sequences of major human coronaviruses.** The sequences of SARS-CoV-2 Wuhan (YP_009724390), MERS-CoV (QFQ59587.1), HCoV-OC43 (QDH43719.1), HCoV-NL63 (AKT07952), HCoV-229E (AOG74783.1) and HCoV-HKU1 (YP_173238) were retrieved from NCBI database. The sequences of B.1.1.7, D614G, B.1.351, B.1.525, B.1.617 (Delta), P.1(B.1.1.281) and B.1.1.529 (Omicron) were obtained from Global Initiative on Sharing Avian Influenza Data (GISAID). Boxed fragments represent IEDB predicted linear antibody epitope (http://tools.iedb.org/bcell/).

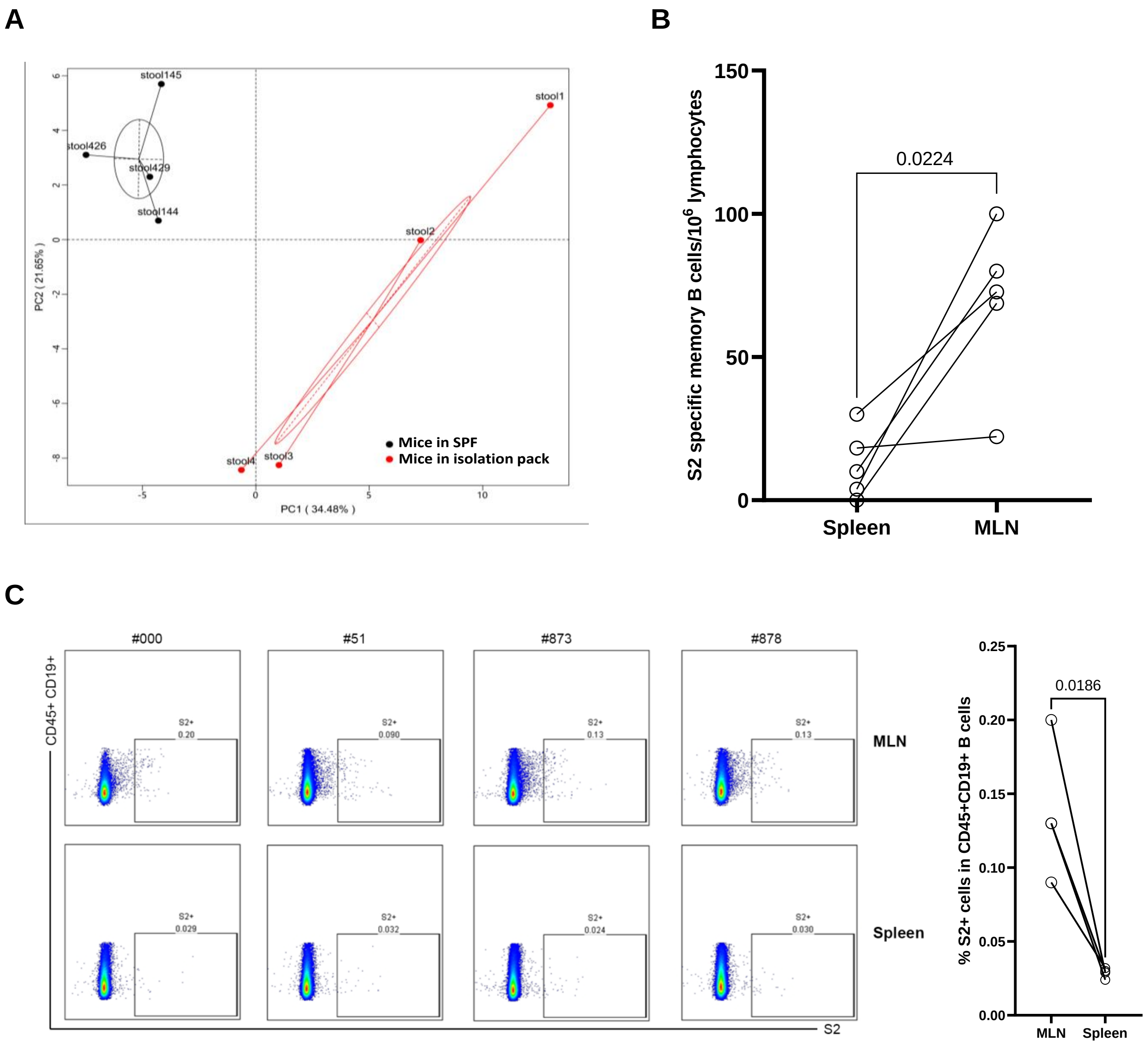


**Supplementary Figure 3 Evidences implied that pre-existing S2 cross-reactive antibodies were associated with commensal gut bacteria**. (**A**) Principle component analysis (PCA) of gut microbial communities between 4 mice in SPF condition and 4 mice in a sterile isolation pack. (**B**) S2 specific memory B cells were measured using the method of memory B cell ELISPOT assay (Cat#3825-2A, Mabtech, Sweden), n=5. Briefly, lymphocytes isolated from mouse MLN or spleen were stimulated with R848 and IL-2. 72 hours later, the cells were washed with R10 and then added into a 96-well ELISPOT plate coated with 10μg/ml S2 protein. After being incubated at 37℃ for 20 hours, the plate was washed with PBS, detecting antibody and Streptavidin-Alkaline Phosphatase were added sequentially. Finally, the plate was developed using BCIP/NBT-plus Substrate (Cat#3650-10, Mabtech, Sweden). Spots were enumerated using an automated ELISPOT plate reader. Statistical analysis was performed by the method of paired t-test. (**C**) Lymphocytes were isolated from MLNs and spleens of 4 naive C57BL/6J mice with relatively high levels of pre-existing S2 reactive antibodies, n=4. S2 specific B cells were defined as CD45$^+$CD19$^+$S2$^+$ (Left). Statistical analysis was performed by the method of paired t-test (Right).

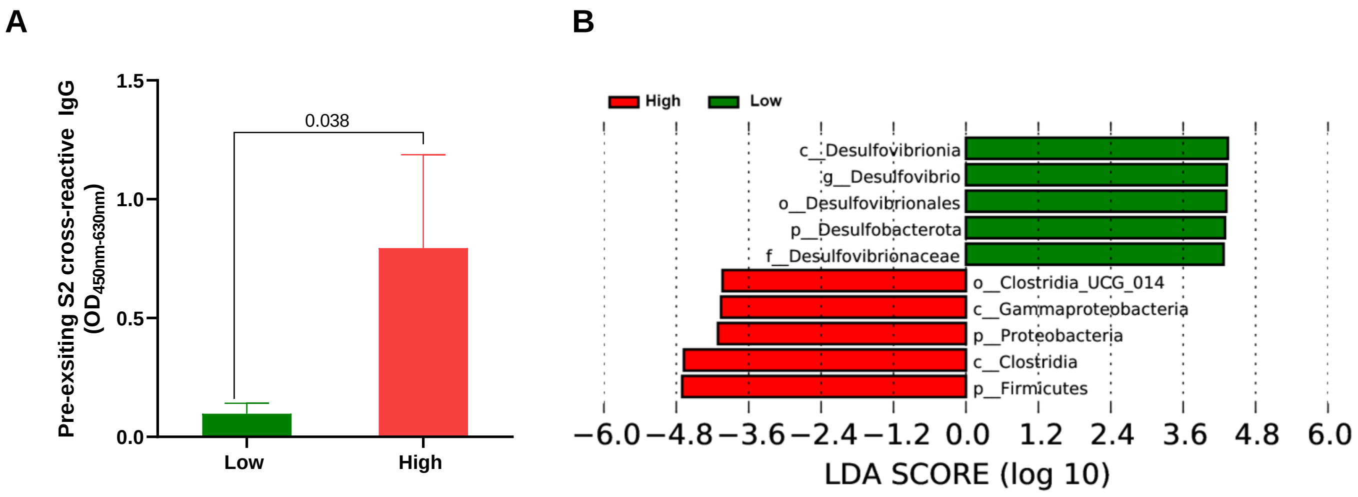


**Supplementary Figure 4 Comparison of gut microbiota composition between mice with low or high levels pre-existing S2 reactive antibodies.** Peripheral blood and stool samples were collected from 3 mice with high levels of pre-existing S2 reactive antibodies and 3 mice with low levels of pre-existing S2 reactive antibodies. All the mice were housed in cages different from each other. (**A**) The levels of pre-existing S2 reactive antibodies were compared the two groups. (**B**) Commensal gut bacteria compositions were analyzed by 16S rDNA sequencing and compared between the two groups. Bacterial abundances were compared by linear discriminant analysis (LDA) analysis and shown as the histogram of LDA scores. Data in (**A**) were shown as mean±SD, n=3. Statistical analysis for (**A**) was performed by the method of unpaired t-test.

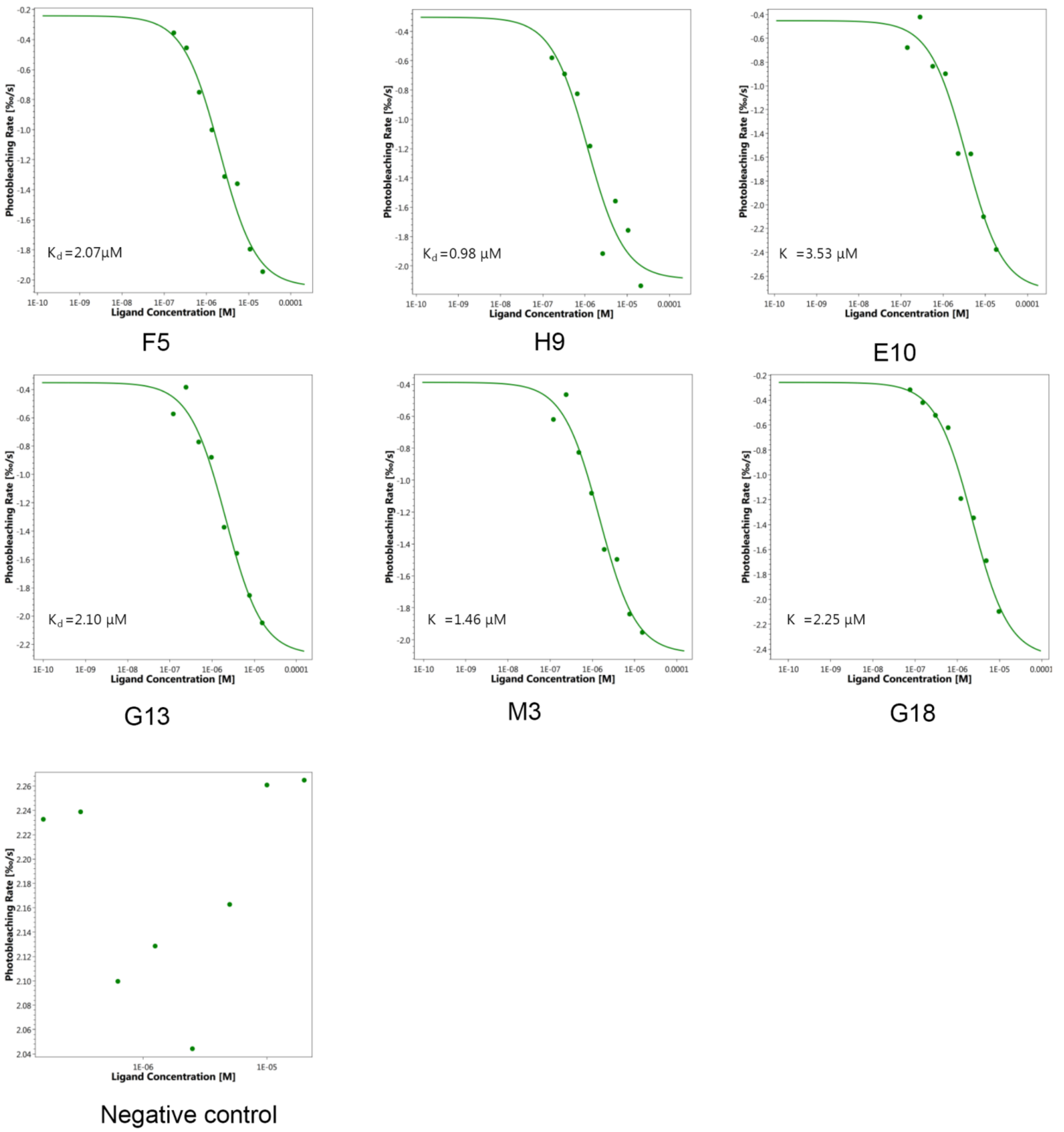


**Supplementary Figure 5 The binding affinities of the P144 reactive mAbs to S2 protein.** The affinities were measured using MicroScale Thermophoresis (Monolith NT.115, Nanotemper, Germany) following the manufacturer`s instruction. Briefly, purified S2 protein (Cat# 40590-V08B，Sino Biological, China) was labeled with the fluorescent dye Red-NHS using Monolith NT protein labeling kit (#MO-L011, Nanotemper, Germany). NHS-Red labeled S2 was incubated with serially diluted mAbs at a final concentration of 5 nM in PBS-T buffer. Five minutes later, the samples were loaded into glass capillaries (Cat#MO-K002, Nanotemper, Germany ) and affinity measurements were performed at 37℃. Dose-response curves of 6 mAbs were acquired and the Kd values were calculated by the MO. Affinity software. Uncoupled NHS-Red dye was used as a negative control.

**A**

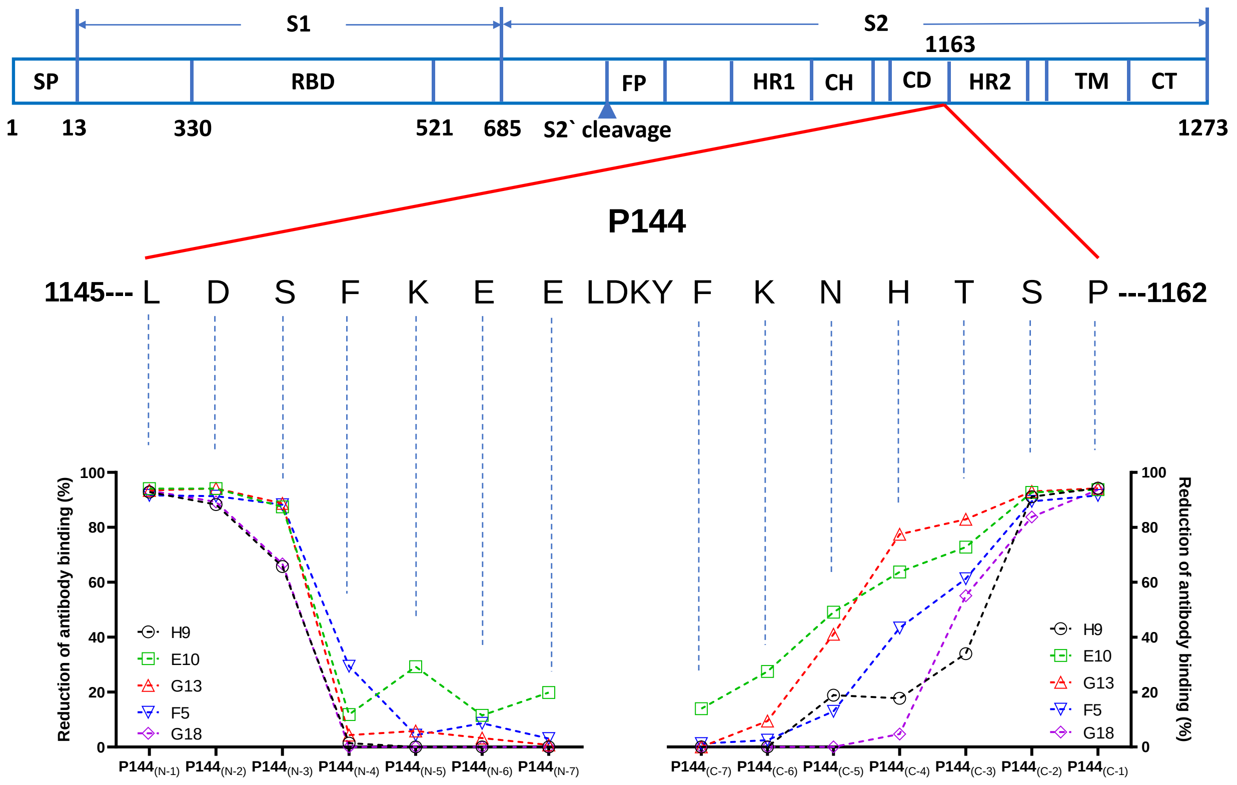


**B**

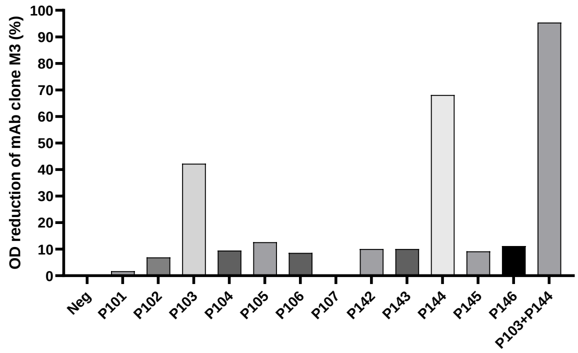


**P144: 1145- LDSFKEELDKYFKNHTSP-1162**

**P103: 817- FIEDLLFNKVTLADAGFI-834**

**Supplementary Figure 6 Characterizations of the minimal epitope recognitions for the 6 monoclonal antibodies isolated from naïve mice**. Six P144 specific monoclonal antibodies were isolated from 2 naïve SPF mice using the hybridoma technology. (**A**) The minimum epitope recognitions of clones H9, E10, G13, F5 and G18 were similar detected by a method of competitive ELISA. Purified S2 protein was used as the coating antigen and the truncated peptides derived from P144 were used as the competitors. (**B**) The epitope recognition of clone M3 was analyzed using purified S2 protein as the coating antigen and each listed peptide was used as the competitor. SP, signal peptide; RBD, receptor-binding domain; FP, fusion peptide; HR, heptad repeat; CH, central helix; CD, connector domain; TM, transmembrane domain; CT, cytoplasmic tail.

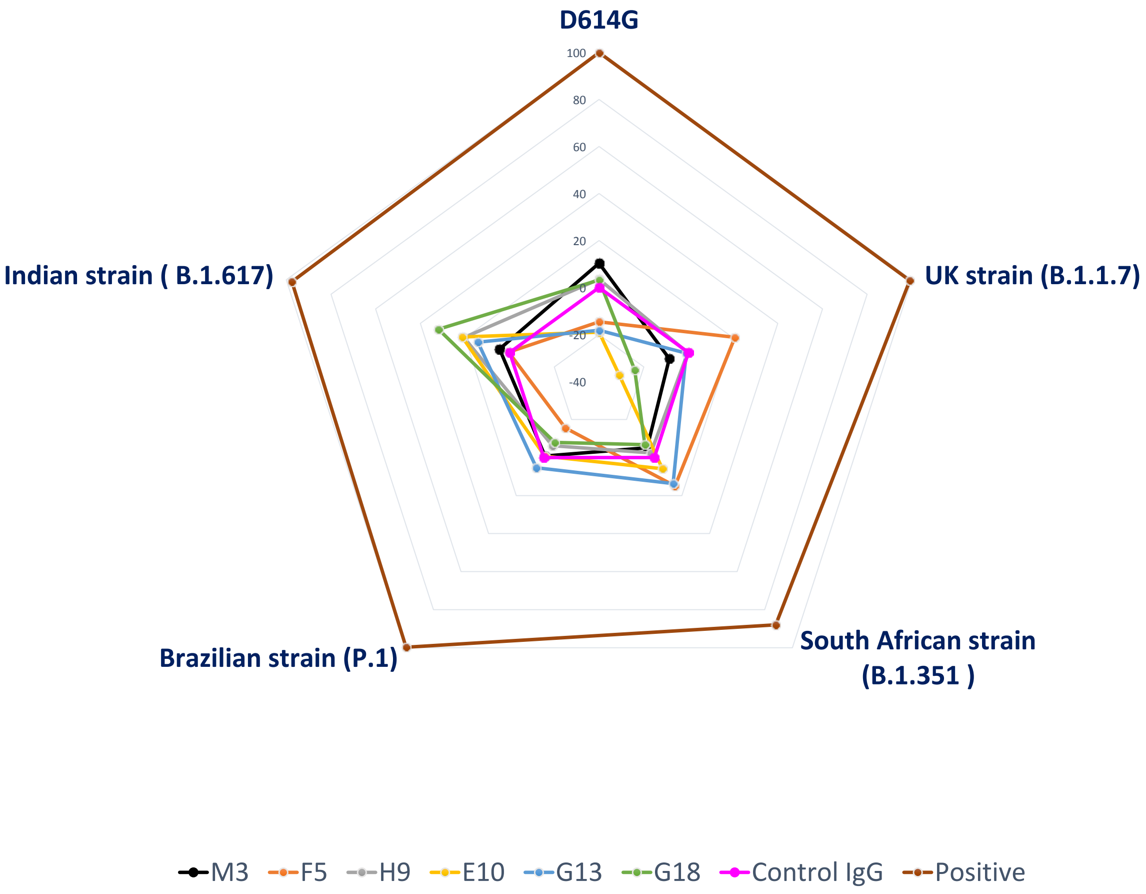


**Supplementary Figure 7 Neutralizing activities of the purified mAbs as measured using a pseudo virus neutralization assay.** Neutralization activities of 6 isolated mAbs against 5 SARS-CoV-2 variants (D614G, B.1.617, B.1.1.7, B.1.351 and P.1) were examined using a pseudo virus neutralization assay. Purified mouse IgG was used as a negative control and serum from an RBD protein vaccinated goat was use as a positive control. Purified mAbs and control mouse IgG were tested at a concentration of 1μg/ml; positive goat serum was diluted at 1:90.

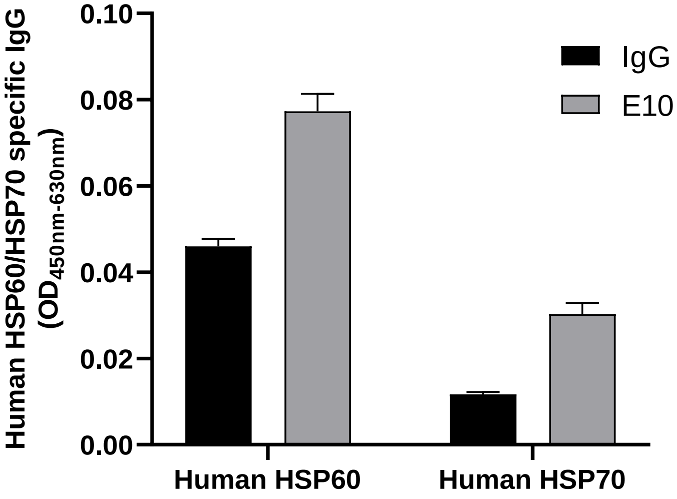


**Supplementary Figure 8 Recognition of the human HSP60 and HSP70 proteins by a P144 reactive mAb (E10).** Purified human HSP60 and HSP70 proteins were used as the coating antigens. E10 or a purified mouse IgG was used as primary antibody at the concentration of 10μg/ml.

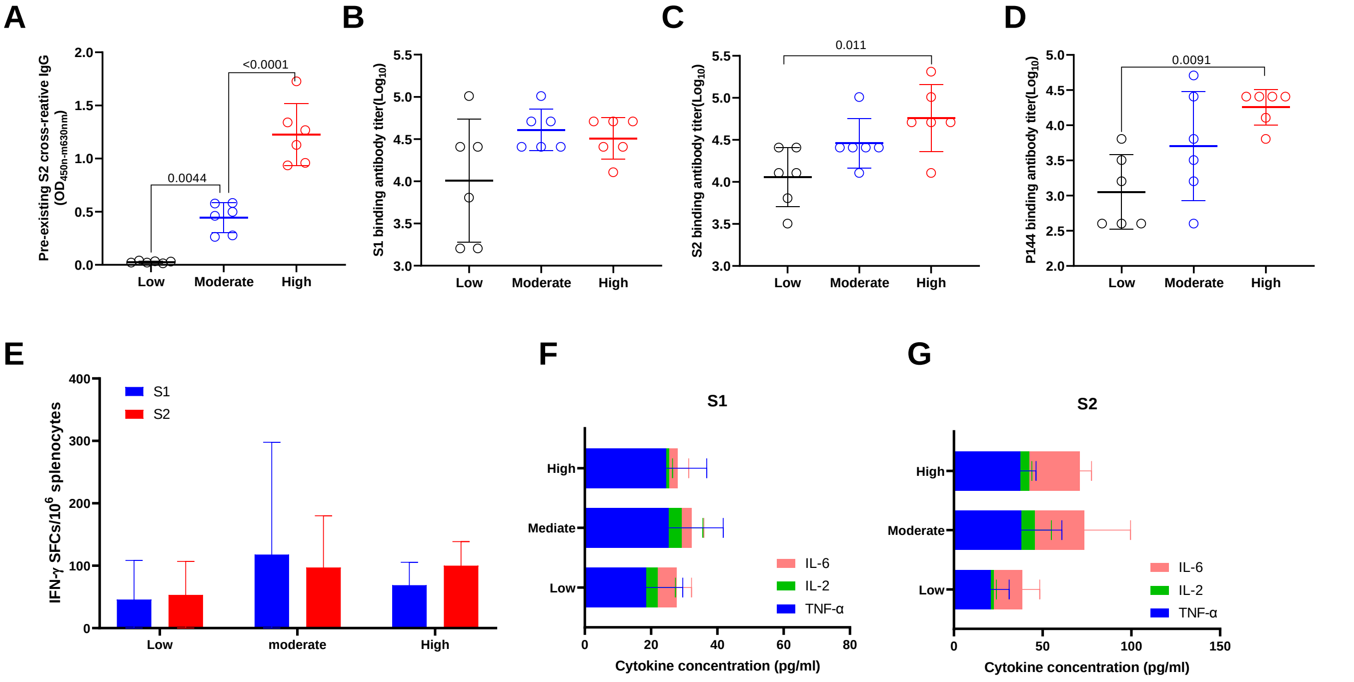


**Supplementary Figure 9 Impact of pre-existing antibodies on the immune responses elicited by a DNA vaccine encoding SARS-CoV-2 S protein (Validating experiment).** 50μg of the DNA vaccine was injected intra muscularly into each mice at week 0, week 2 and week 4, respectively. Two weeks after the final vaccination, the mice was euthanized for the measurements of specific immune responses. (**A**) Peripheral blood was collected before immunization and levels of pre-existing S2 specific IgG were compared among three groups. (**B**) Comparisons of endpoint IgG titers against S1 measured at 2 weeks post the final vaccination. (**C**) Comparisons of endpoint IgG titers against S2 measured at 2 weeks post the last immunization. (**D**) Comparisons of P144 specific IgG titers measured using BSA-P144 conjugate as the coating antigen (**E**) S1 and S2 specific IFN-γ responses were compared among different groups at 2 weeks post the final immunization. S1 (**F**) and S2 (**G**) specific releases of IL-2, IL-6 and TNF-α as measured using the method of multiplex cytokine bead assay were also compared among different groups. Data were shown as mean± SD, n=6. SFCs, spot forming cells. Statistical analyses were performed by the method of one-way ANOVA.

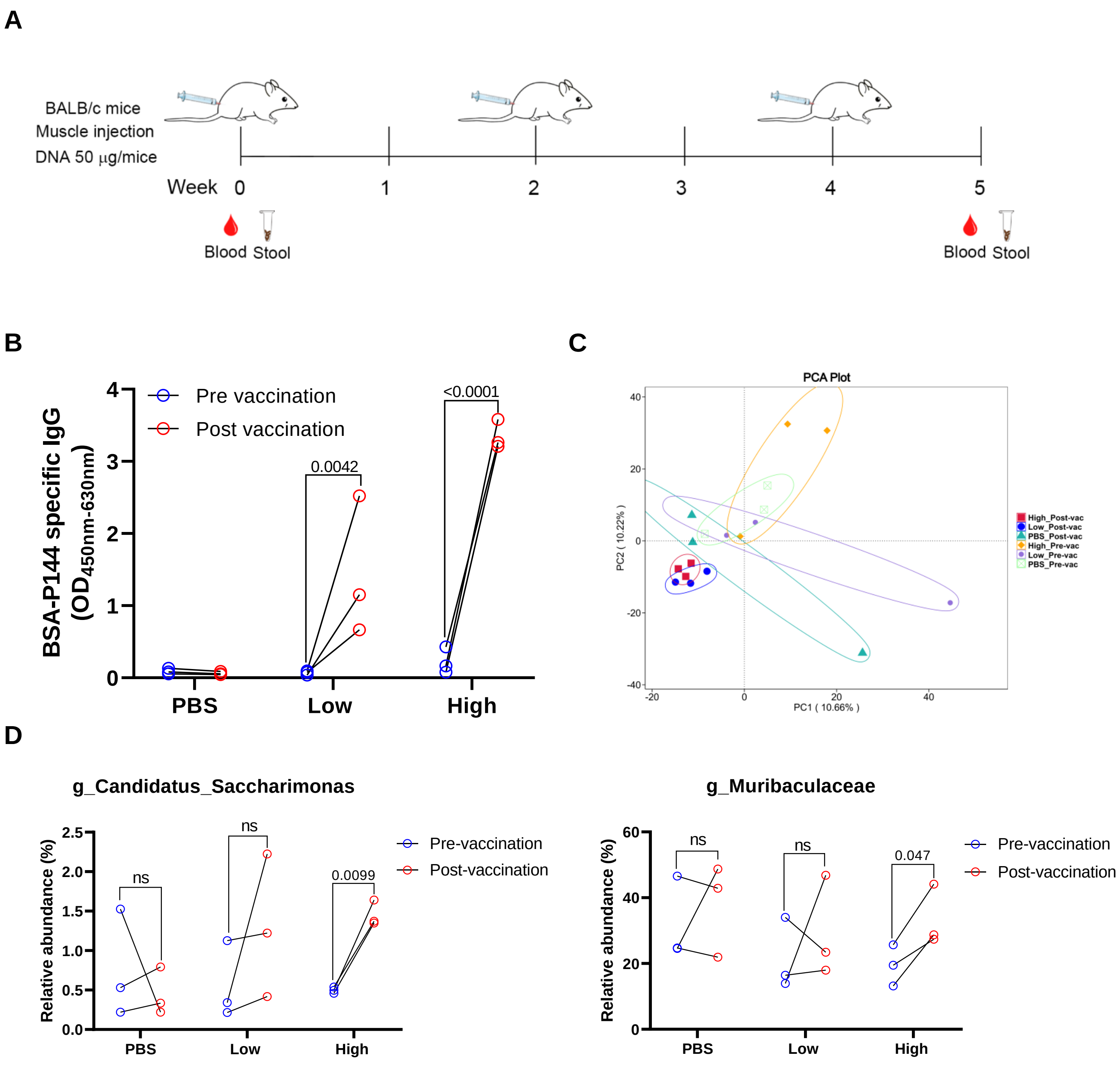


**Supplementary Figure 10 The impact of SARS-CoV-2 S DNA vaccination on the gut microbiota composition of mice. (A)** Schematic illustration of the experiment schedule. 50μg of the DNA vaccine was injected intra muscularly into each mice at week 0, week 2 and week 4, respectively. Stool samples were collected at baseline and 5 weeks after the last vaccination. (**B**) Comparisons of P144 reactive antibody levels between mouse sera collected before and after vaccination. (**C**) Principle component analysis (PCA) of gut microbiota composition. (**D**) Comparisons of relative abundances of representative bacteria genus that were found to be impacted by vaccination. High, group of mice with high levels of pre-existing P144 reactive antibody; Low, group of mice with low levels of pre-existing P144 reactive antibody. Statistical analyses were performed by the method of paired t-test.

#### Supplementary Table 1 V(D)J gene sequencing of P144 reactive mAbs

| mAb Clone No. | Chain type | Top V gene match | Top D gene match | Top J gene match | CDR1 | CDR2 | CDR3 |
|---|---|---|---|---|---|---|---|
| E10 | VH | IGHV5-9*02 | IGHD2-3*01 | IGHJ2*01 | GFAFSSYD | ISSGGSYT | ARQDGYYRYFDY |
| | VL | IGKV8-30*01 | | IGKJ1*01 | QSLLYSSNQKNY | WAS | QQYYSYPPT |
| F5 | VH | IGHV2-5*01 | IGHD1-1*01 | IGHJ2*01 | GFSLTSYG | IWRGGST | AKIDGSSNY |
| | VK | IGKV6-23*01 | | IGKJ2*01 | QDVGTA | WAS | QQYSSYPT |
| G13 | VH | IGHV2-5*01 | IGHD1-1*01 | IGHJ2*01 | GFSLTSYG | IWRGGST | AKIDGSSNY |
| | VL | IGLV2*02 | | IGLJ2*01 | TGAVTTSNY | GTS | ALWYSTHYV |
| G18 | VH | IGHV2-5*01 | IGHD1-1*01 | IGHJ2*01 | GFSLTSYG | IWRGGST | AKIDGSSNY |
| | VK | IGKV6-23*01 | | IGKJ2*01 | QDVGTA | WAS | QQYSSYPT |
| H9 | VH | IGHV2-5*01 | IGHD1-1*01 | IGHJ2*01 | GFSLTSYG | IWRGGST | AKIDGSSNY |
| | VK | IGKV6-23*01 | | IGKJ2*01 | QDVGTA | WAS | QQYSSYPT |
| M3 | VH | IGHV2-5*01 | IGHD1-1*01 | IGHJ2*01 | GFSLTSYG | IWRGGST | AKIDGSSNY |
| | VK | IGKV6-23*01 | | IGKJ2*01 | QDVGTA | WAS | QQYSSYPT |

**Supplementary Table 2 Pre and post vaccination antibody responses of 28 healthy vaccinees**

| NO. | Post vaccination Neutralizing antibodies (ng/ml) | Post vaccination RBD binding antibody titer (Log) | Pre-existing S2 binding antibody titer (Log)♠ | Pre-existing BSA-P144 binding antibody titer (Log)♣ |
|---|---|---|---|---|
| C2 | 0 | 2.431363764 | 2.903089987 | 2.30103 |
| C3 | 0 | 1.954242509 | 2.602059991 | 2 |
| C4 | 0 | 2.431363764 | 3.505149978 | 2.30103 |
| C5 | 0 | 1.954242509 | 2.301029996 | 2 |
| C7 | 0 | 1.954242509 | 2.301029996 | 2 |
| C8 | 7.692307692 | 1.954242509 | 2.602059991 | 2 |
| C9 | 190.8215385 | 2.908485019 | 2.903089987 | 2 |
| C10 | 0 | 1.954242509 | 2.301029996 | 2 |
| C11 | 819.6161538 | 3.385606274 | 3.806179974 | 2.30103 |
| C12 | 25.00923077 | 1.954242509 | 2.602059991 | 2 |
| C14 | 41.14384615 | 1.954242509 | 2.602059991 | 2 |
| C15 | 252.3284615 | 3.385606274 | 2.301029996 | 2 |
| C16 | 819.9984615 | 3.385606274 | 2.602059991 | 2 |
| C17 | 0 | 1.954242509 | 2.602059991 | 2.30103 |
| C18 | 49.34769231 | 1.954242509 | 2.301029996 | 2 |
| C19 | 46.23615385 | 2.431363764 | 2.602059991 | 2 |
| C20 | 82.89076923 | 2.431363764 | 2.301029996 | 2 |
| C23 | 64.88846154 | 2.431363764 | 3.505149978 | 2 |
| C24 | 96.24692308 | 2.908485019 | 3.505149978 | 2.60206 |
| C25 | 76.72846154 | 2.908485019 | 2.602059991 | 2.30103 |
| C26 | 74.09615385 | 2.908485019 | 2.903089987 | 2.30103 |
| C27 | 43.59692308 | 2.431363764 | 2.602059991 | 2.30103 |
| C28 | 0 | 1.954242509 | 2.301029996 | 2 |
| C30 | 305.9192308 | 3.385606274 | 2.903089987 | 2.60206 |
| C31 | 125.2430769 | 2.908485019 | 2.602059991 | 2.60206 |
| C32 | 30.08615385 | 2.908485019 | 3.505149978 | 2.60206 |
| C35 | 173.7146154 | 2.908485019 | 2.301029996 | 2 |
| C36 | 318.0553846 | 3.862727528 | 2.903089987 | 2.30103 |

Note: Heat denatured human sera were used as negative control. "♠", Pre-existing S2 binding antibody titers positively correlated with the post vaccination RBD binding antibody titers (r=0.468, p=0.012). "♣", Pre-existing BSA-P144 binding antibody titers positively correlated with the post vaccination RBD binding antibody titers (r=0.497, p=0.007). Statistical analyses were performed using the method of Spearman's correlation.
